# Associations between sickle haemoglobin and the *Plasmodium falciparum CLAG* and *FIKK* gene families revealed by meta-analysis of 6,289 African samples

**DOI:** 10.1101/2025.11.18.25340465

**Authors:** Annie J. Forster, James Docker, Katie Healey, Zhipeng Zhang, Amy Trebes, David Buck, William L. Hamilton, Lucas Amenga-Etego, Alexander J. Mentzer, Alfred Amambua-Ngwa, Gavin Band

## Abstract

The recent discovery that mutations in the *Plasmodium falciparum* genome are overrepresented in infections of sickle haemoglobin (HbS)-carriers has highlighted new questions about the underlying biological and evolutionary interaction, yet the full extent of this association is unknown. By meta-analysing host and parasite data from N=6,289 infections, including 831 newly sequenced samples from The Gambia, we implicate several new parasite genome regions in the interaction, including within the *CLAG3* nutrient uptake channel and the threonine/serine kinase *FIKK3* on chromosome 3. The HbS-associated mutations share unusually strong linkage disequilibrium, and we use a series of analyses to disentangle their complex genetic structure and independent effects. The most prominent signal is observed at a polymorphism shared between two *CLAG3* paralogs, estimated to halve the level of protection due to HbS. Alongside previous findings in *ACS8* and *FIKK4*.*2*, these results now implicate three of the major *P. falciparum* gene families in host-parasite interactions and open new avenues for functional inquiry.

## Introduction

Humans have evolved multiple resistance mechanisms to counter malaria infection^1,2^. Among these, sickle haemoglobin (HbS), encoded by a point mutation in the beta-globin gene on chromosome 11 (rs334 A>T), provides the strongest known protection against severe *Plasmodium falciparum* malaria ^1^. Over decades of study, HbS has become a model of host resistance to malaria, and several biological mechanisms have been proposed to explain its protective effect^3^.

However, recent findings have also suggested that parasites may be evolving in response to HbS resistance. The two genetic discovery studies conducted so far were an analysis of severe infections from The Gambia and Kenya ^4^ and an analysis of mild infections from Ghana^5^, which revealed that mutations in four regions of the parasite genome (termed *P. falciparum* sickle-associated (*Pfsa+*)1-4) are over-represented in infections of HbS carriers. The estimated protection conferred by HbS was substantially reduced against parasites carrying these mutations^4^. These findings have also been supported by a study in asymptomatic infections in Cameroon^6^ and by population-level association^7^, and collectively suggest the variants have evolved to overcome the protective effects of HbS.

There is great interest in understanding the biological and evolutionary impact of the *Pfsa+* mutations, whose analysis is complicated by several features including between-locus linkage disequilibrium and population-specificity of some of the loci^4,5^. However, given the limited number of HbS genotype individuals contained in current datasets, it is also possible that the full extent of the HbS association signal may not yet have been discovered. To address this, we here generate new parasite whole genome and host HbS genotype data from a set of *N*=831 severe and non-severe infections collected in The Gambia in 1988-90. First, we use these to conduct a parasite genome-wide association for severity. We then combine this with the existing datasets to yield a resource of more than 6,000 infections with both parasite and human genetic data and use this to meta-analyse HbS-*Pfsa* associations across Africa. Our analysis reveals multiple previously unreported signals of association with HbS in several genome regions, including within the *CLAG* and *FIKK* gene families, and we implement a series of methods to analyse these associations in detail accounting for the complex observed paralogy, linkage disequilbrium, and locus structural variation.

## Results

### Generating a new resource to investigate HbS-*Pfsa* relationships

We first generated whole-genome variation data for a set of 831 samples from The Gambia. These were collected as part of an epidemiological study at the Royal Victoria Teaching Hospital during periods of high malaria endemicity between 1988-1990 and were the basis of a previously reported association between human leukocyte antigen (HLA)-B*53 and protection against severe disease^8^. This dataset, which we refer to as the ‘Genetic Analysis of Malaria Cases and Controls’ (GAMCC) resource, is of interest because it dates from a timepoint before the widespread use of chloroquine in The Gambia and thus expands the temporal coverage of data in this region (**Fig. 1A**). A subset of 558 samples were ascertained with severe symptoms, while the remainder had elevated parasitaemia but without severe complications (referred to as non-severe below) (**Methods)**.

**Figure 1.**
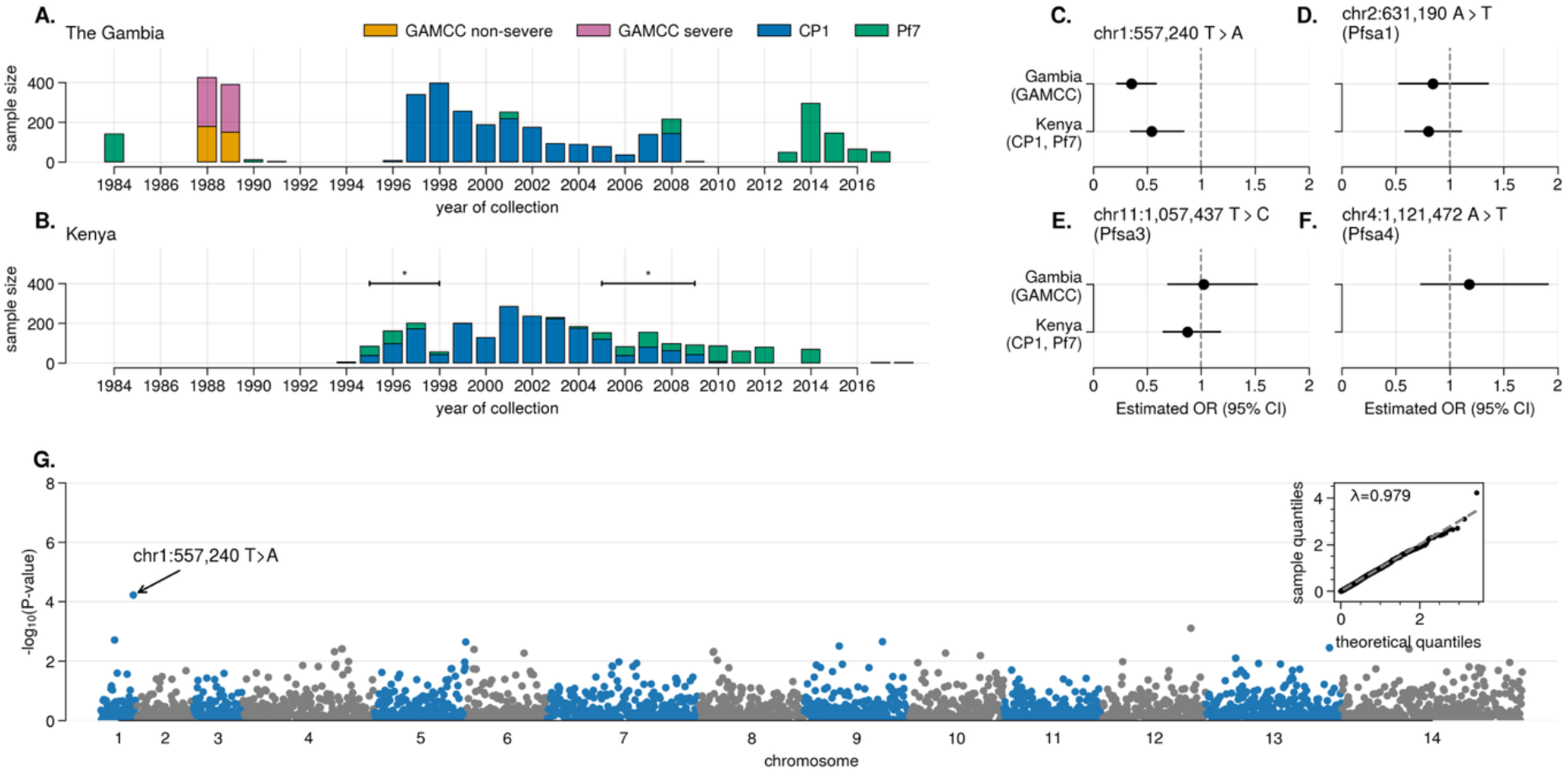
Details of the GAMCC resource and evidence for parasite genetic effects on disease severity. Details of the collection timepoints, sample sizes and disease status across multiple datasets in **A**. The Gambia, including GAMCC (yellow) and **B**. Kenya as indicated by the legend in panel A. **C-F**. Forest plot showing estimated effect size for the chr1:557,240 A>T variant, and previously reported *Pfsa1, 3*, and *4* variants, on severe versus non-severe disease in GAMCC, as well as in a comparison of severe (CP1) and non-severe (Pf7) infections from coastal Kenya (using samples collected over a shared time frame as indicated by a * on panel B; see **Supplementary Info). G**. Evidence for association (-log10(P-value), y-axis) between *P. falciparum* genetic variants (points) across the genome (x-axis) with severe vs. non-severe disease status in the GAMCC data. To avoid spurious effects due to the relatively small sample, variants with minor allele frequency < 7.5% were excluded from this analysis. A quantile-quantile plot for this test and inflation factor lambda (λ) is also shown.

To generate parasite sequencing in these samples, we used selective whole-genome amplification of parasite DNA followed by sequencing samples on the Illumina NovaSeq platform. We then called genetic variants jointly with the Malaria Genomic Epidemiology Network (MalariaGEN) CP1^4^ samples using a GATK HaplotypeCaller pipeline^9^ (full details in **Methods)**. For the analyses below, we focus on biallelic variants in the core genome (i.e. excluding sub-telomeric and other complex regions where variation is challenging to call using short-read sequencing data^10^). Host typing at the HbS locus was carried out using a combination of approaches, with most samples determined by Sanger sequencing (**Methods)**.

The GAMCC dataset provides a key opportunity to test whether parasite genetic variation is associated with disease severity. We began by conducting a genome-wide analysis of severe vs. non-severe in these samples (**Fig. 1G**), revealing some evidence for association at a single variant on chromosome 1 (chr1:557,240 A>T; odds ratio (*OR*)=0.36, 95% confidence interval (CI)=0.22–0.59; P-value (*P*)=6.1×10^−5^) (**Fig. 1C** and **Supplementary Info**). The T allele at this site is at about 15% frequency in non-severe cases and 6% in severe cases in this sample (**Fig. 1B**). The variant lies 1kb downstream of an exported protein (*PF3D7_0114500, hyp10*) and approximately 1.8kb upstream of an annotated erythrocyte membrane protein pseudogene (*PF3D7_0114400, VAR*) (**Figure S2**).

Although available data to replicate this signal is currently limited, a comparison of the frequency of this allele in two sample sets from Kilifi County in Kenya, collected in overlapping time periods (between 1995-98 and 2005-09 (**Fig. 1B**)) also showed a similarly reduced frequency in severe infections compared to population samples (**Fig. 1C**, *f*^P*f*7^=9.6%, *f*^CP1^=5.4%, *P*=0.015, **Supplementary Info**). This provides some evidence that parasite genetic variation associates with disease severity, but we caution that this analysis is constrained by the relatively small sample size of the GAMCC collection.

We also specifically inspected the previously reported HbS-associated variants in this comparison. No strong evidence of a difference in *Pfsa+* allele frequency was observed between severe and non-severe cases, at either the *Pfsa*1, *Pfsa3*, or *Pfsa4* loci (**Figs. 1D-F**, *P* > 0.45). The *Pfsa2+* allele (chr2: 814,288 C>T) is largely absent from west Africa and is not observed in the GAMCC dataset. These results therefore suggest that the association with HbS does not substantially differ between severe and non-severe cases in this set. For our main analysis below, we analyse these phenotypes in combination.

### Joint analysis of HbS associations using cross-population data

The GAMCC dataset adds to four others which contain both parasite genome-wide genotyping and host genotyping for the HbS allele. These are: the MalariaGEN set of severe malaria cases from The Gambia and Kenya (CP1) collected from infants in 1995-2009 (**Figs. 1A & 1B**) ^4^; a set of mild malaria infections within the MalariaGEN Pf7 resource collected in 2015-2018 in Ghana^5^; and a set of RNA-seq data collected from 16 HbAS and 16 HbAA children with mild infections from Mali^11^ (**Fig. S1)**. The total sample size across datasets is 6,289 including 112 HbAS and 14 HbSS individuals, and 342 with HbC genotypes (of 6,079 typed samples) (**Supplementary Table 1**).

To test for association between HbS and *P. falciparum* genetic variation, we used a logistic regression approach implemented in HPTEST^4^, working separately within each of the five component datasets. For our main analysis we grouped the HbAS and SS genotypes together (i.e. assuming dominance) and treated parasite genotype as the outcome variable. For simplicity, samples with mixed genotype calls at each variant were excluded. We then used the BINGWA approach ^1^ to combine evidence across datasets under fixed effect and population-variable models of association. The primary measures of evidence in this method are a model-averaged Bayes factor (*BF*_*avg*_) and a fixed-effect meta-analysis *P*-value. Under reasonable assumptions (**Methods**), a *BF*_*avg*_ of around 1×10^6^ may be considered strong evidence for association, and in the following we focus on variants above this threshold which are further detailed in **Supplementary Table 2**.

The previously identified *Pfsa*1 and *Pfsa*3 variants strongly replicated in the GAMCC dataset (**Supplementary Info**). Notably, our analysis also identified several additional loci that have substantial evidence for association with HbS (**Fig. 2-3, Fig. S3** and **Supplementary Table 2**). These include several regions of association on chromosome 3, including nonsynonymous variants in the genes *CLAG3*.*1* and the serine/threonine protein kinase *FIKK*3, as well as a weaker association in the 5’ untranslated region of an EMP1 trafficking protein (*PTP7*) (**Fig. 2C**).

**Figure 2.**
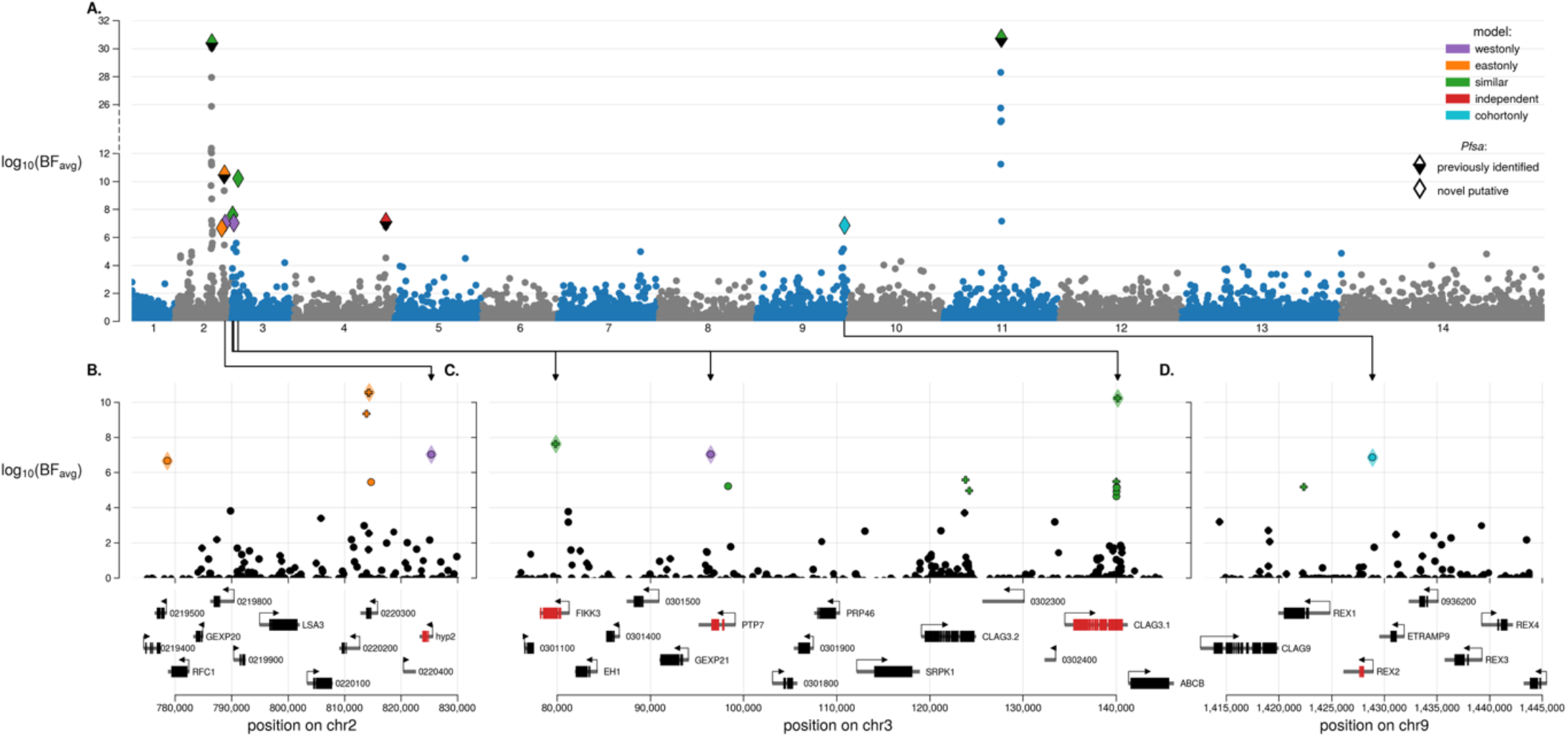
Evidence for association between *P. falciparum* variants and the HbS genotype across five datasets. **A**. Points show evidence for association between *P. falciparum* variants and HbS genotype meta-analysed across five datasets. Each point represents one biallelic variant in the *P. falciparum* genome with at least a 1% allele frequency across the combined dataset. Results were computed by logistic regression within each dataset, using parasite genotype as the outcome variable and HbS genotype as the predictor. No additional covariates were included, and we assumed a dominant model of association i.e. grouping HbAS and HbSS genotypes. Samples with missing genotype calls, and those with mixed *P. falciparum* genotype calls were excluded from the calculation at each variant. The log10 Bayes factor for association averaged across models (*BF*_*avg*_) (y-axis) was then computed by meta-analysis across datasets at each *P. falciparum* variant (x-axis). To capture potential population-specific effects, *BF*_*avg*_ averages over models of association (colours) including: effects similar across populations (‘similar’ model, green); effects independent across populations (‘independent’ model; red); where non-zero effects are restricted to west or east African datasets (“west only”, purple, or “east only” orange); and dataset-specific effects (light blue). Lead variants (those with the highest *BF*_*avg*_ in each region identified with log_10_(*BF*_*avg*_) > 6 in the meta-analysis) are marked with a diamond; to be marked, variants are required to be at least 10,000 bp apart (except at *Pfsa1* and *Pfsa3* where a minimum separation of 15,000 bp was applied). Diamond colour indicates the model with the highest supporting evidence at the lead variant; those previously reported^4^ are partially shaded black. **B-D**. show zoomed views detailing associations at putative *Pfsa* loci, with sites of interest connected to their position in plot A using arrows. In addition to the same diamond markings as subplot A, any variant where log_10_(*BF*_*avg*_) > 4 is coloured by model of greatest *BF*. In these plots a ‘+’ marker indicates the mutation causes a non-synonymous amino acid change within the encoded protein. Below shows the positions of genes in the region, and genes that are highlighted within the main text are marked as red.

We also observed evidence of association at a variant on chromosome 2 in an uncharacterised exported protein (*hyp2*) (**Figs. 2B & 3)**. Although this lies close to the previously reported *Pfsa2+* mutation, the association shows an opposing pattern of geographic support with strongest evidence in west African datasets (**Fig. S4A**). In the Kenyan dataset, a further signal was observed within the ring exported protein 2 (*REX2*) gene on chromosome 9 (**Fig. 2D)**. This variant was relatively rare in Kenya (*f*=3.7%) and was not polymorphic in West African populations so cannot be independently replicated using current data (**Figs. 3 & S4E**).

**Figure 3.**
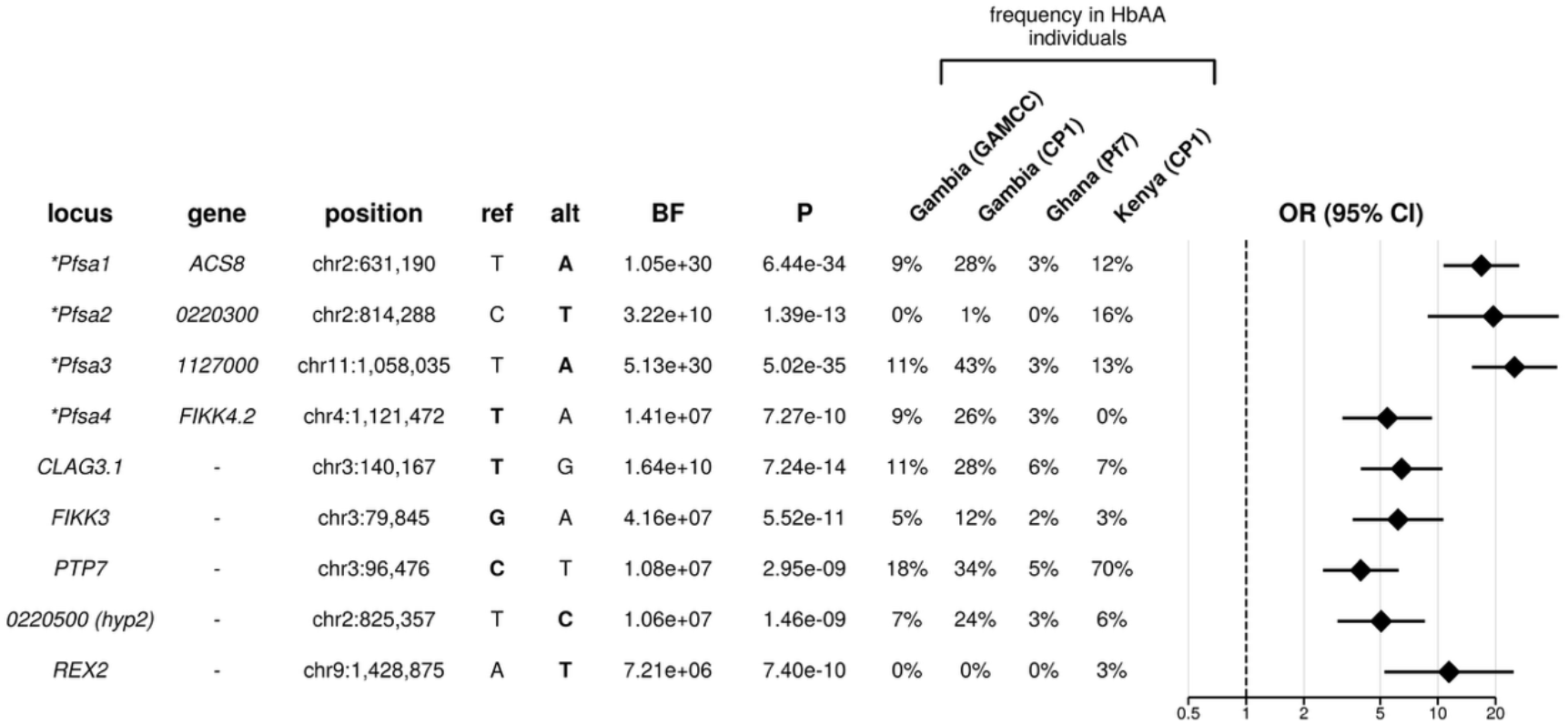
Detail of association between HbS and *P. falciparum* variants with strongest evidence. Points indicate the estimated meta-analysis odds ratio (OR) and 95% confidence interval (CI) for loci with the strongest HbS-associations across the *P. falciparum* genome (log_10_(*BF*_*avg*_) > 6) across five datasets. These include four previously described *Pfsa* loci (*Pfsa1-4*, marked with an asterisk) and five additional putative HbS-associated loci identified in this analysis. To the left of each estimate are the genomic position relative to the Pf3D7_v3 reference genome assembly, reference and alternate alleles (with the HbS-associated allele indicated in bold), Bayes factor (BF), frequentist p-value (P) and frequency of the allele in HbAA individuals in each of the four genomic datasets. ORs are shown with respect to the HbS-associated allele and are further reported in **Supplementary Table 2**.

As outlined above, the association evidence for the newly implicated variants is less pronounced than at the *Pfsa1-3* loci but is nevertheless compelling. Apart from *REX2*, they all have support in multiple datasets with some variation between populations (**Fig. S4)**. The effect sizes of these associations are also considerable (e.g. *CLAG3*.*1*: *OR*=6.67 (4.0-11.11); *FIKK3*: *OR*=6.25 (3.57-11.11); **Fig. 3**) but weaker than those for the previously reported HbS-associated loci (**Supplementary Table 2**).

Taken together, these results suggest that the association between *P. falciparum* genetic variants and HbS is complex and involves more variation than has been described so far. A complication is that the newly associated variants, along with *Pfsa1-4*, show elevated levels of between-locus linkage disequilibrium (LD) (i.e. the genotypes are correlated), despite falling in different genomic regions (**Fig. S5 & S6**). The underlying cause of this LD is unclear but could be due to joint selection of the loci ^7^ or could imply a form of epistatic interaction. The allele frequencies at these loci across Africa are also positively associated with HbS, indicating further evidence for selection (**Fig. S4)**. Across time in The Gambia (**Fig. 1A**), the variants also all appear to have increased in frequency somewhat since the 1960s (**Fig. S7**)^12^.

### Analysis of effects jointly across *Pfsa* loci

The large number of implicated loci raises the question of how they combine to modify infection risk. To assess this, we fit a joint model of association to estimate the contribution of each locus to the relative risk conferred by HbS (estimated using the ratio of relative risks (*RRR*) which compares the protective effect of HbS for each combined parasite genotype relative to *Pfsa*-parasites). In the following, we use “*Pfsa+”* and “*Pfsa-”* to refer to the HbS-associated and non-HbS-associated allele at each of the loci, as previously^4^. We first assessed each locus jointly with *Pfsa1* and *Pfsa3* (**Fig. S8**). Among the HbS-associations which replicated across multiple independent datasets, the *Pfsa+* mutation in *CLAG3*.*1* showed the largest additional effect, approximately doubling the *RR* (estimated fold change in relative risk = 1.97; 95% CI=1.06 − 3.65. It similarly markedly increased the relative risk due to HbS across multiple common *Pfsa1-4* backgrounds (e.g. *RRR*=50.5 vs. *RRR*=7.05 in infections carrying *Pfsa*1+ and *Pfsa*3+; *RRR*=49.5 vs RRR=20.1 in infections carrying *Pfsa1+, Pfsa2+* and *Pfsa3+;* **Fig. 4**). Thus, this analysis provides firm evidence that the *CLAG3*.*1* mutation has a specific association effect that is not simply due to LD with the previously identified loci.

**Figure 4.**
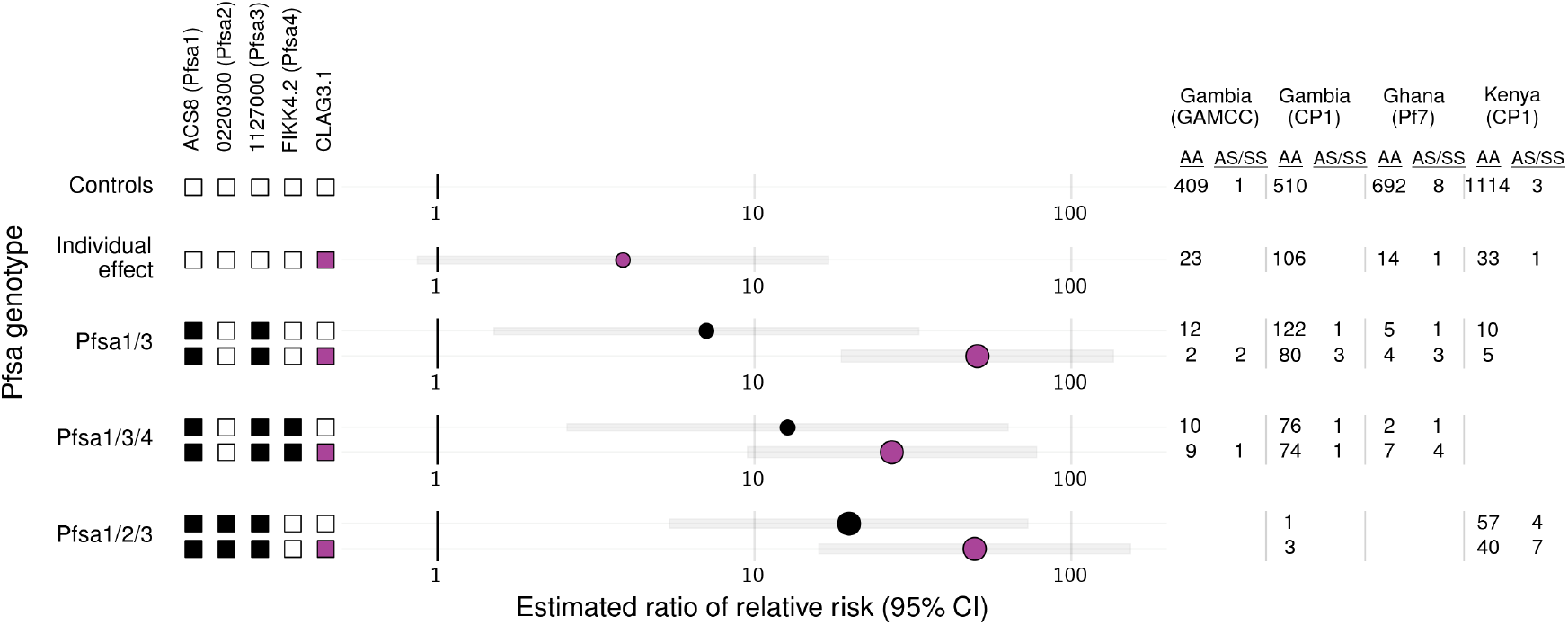
Mutations in the *CLAG3*.*1* gene increase the relative risk of HbS. Estimated ratio of relative risk (RRR) of malaria based on HbS for different *Pfsa* genotype combinations (indicated by filled black squares on the left) compared to infections of *Pfsa-*parasites. This analysis was conducted across all available whole genome sequence data with paired HbS genotyping (GAMCC, CP1 Gambian and Kenyan samples and Ghanaian samples) using genotypes at the previously described *Pfsa1* (chr2:621,190 A>T in the ACS8 gene), *Pfsa2* (chr2:814,288 C>T (0220300)), *Pfsa3* (chr11:1,058,035 T>A (1127000)) and *Pfsa4* (chr4:1,121,472 A>T(FIKK4.2)) loci with the addition of the putative novel *Pfsa+* mutation in the CLAG3.1 gene (chr3:140,167 T>G, highlighted in purple). RRRs were calculated using a multinomial logistic regression using *Pfsa-*infections as a baseline. 95% confidence intervals are indicated by the light grey bar. To avoid over-fitting of points with small sample sizes, Bayesian regularisation was performed using a weakly informative Gaussian prior with mean 0 and standard deviation of 10 for each parameter, and between-parameter correlation set to 0. HbAS and HbSS genotypes were grouped together (i.e. a model of dominance was assumed), and estimates supported by four or more HbAS/SS individuals (and thus having higher confidence), are indicated by larger points. To the right, sample sizes stratified by human genotype are shown per dataset.

The *REX2* mutation also shows a clear effect in this analysis (**Fig. S8**), but as described above is only present in Kenya and will require further replication. For the other loci implicated above, the evidence that the loci independently affect the relative risk due to HbS is more limited. However, we note that larger samples containing more HbS carriers will be required to clarify these estimates; it is also possible that these variants play an indirect role in overcoming HbS resistance which is not captured by this analysis.

### The HbS-associated mutations in *CLAG3* are para-polymorphic

Motivated by the clear signal at *CLAG3*.*1*, we investigated this locus in further detail. *CLAG3*.*1* lies ∼10Kb upstream of a paralogous gene *CLAG3*.*2* on chromosome 3 (**Fig. 2C)**, and both are members of the larger *CLAG* gene family which varies in size across the clade of ape-infecting malaria species related to *P. falciparum*^13^. The *CLAG3* genes encode a component of the Plasmodial surface anion channel (PSAC), which facilitates solute uptake from the host cell environment^14^, and have the highly unusual property that they are mutually exclusively expressed (with only one copy transcribed at any given time)^15,16^. However, it has also been reported that some parasites carry a single *CLAG3* gene (a hybrid known as *CLAG3h*) which arises through a ∼10kb deletion of part of the chromosome^17^ (depicted in **Fig. 5A**).

**Figure 5.**
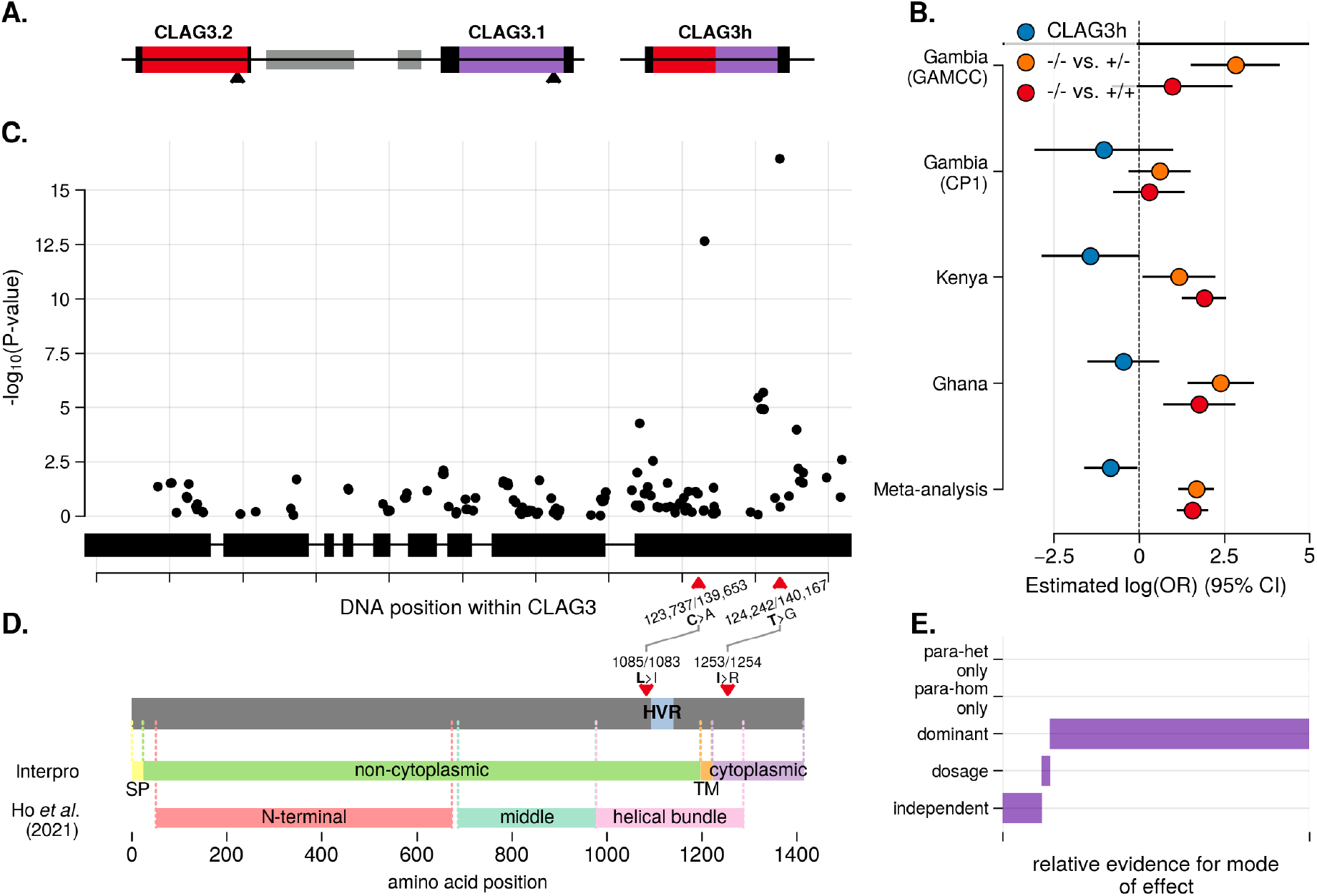
Paralogy-aware evidence for association with HbS across the CLAG3 gene. **A**. A cartoon diagram of the CLAG3 locus. Parasites typically carry a CLAG3.2 (red) and CLAG3.1 (purple) gene on chromosome 3, separated by two pseudogenes (grey) (left). Some parasites instead carry a hybrid gene copy (CLAG3h) due to a ∼10kb deletion. Untranslated regions (black) differ between CLAG3.1 and CLAG3.2, but otherwise paralogues share high levels of sequence identity. Black arrows point to chr3:124,242 T>G (CLAG3.2) and chr3:140,167 T>G (CLAG3.1), a para-polymorphism shared between gene copies. **B**. Forest plots showing estimated association between HbS and the *CLAG3h* deletion (blue) and para-polymorphic genotype (red/orange) at the chr3:124,242/140,167 locus. Results are expressed with respect to the para-homozygous G/G (i.e. *Pfsa* -/-) baseline and reflect T/G (-/+, orange) and T/T (+/+, red). **C**. Points show evidence for association (log10P-value) between *P. falciparum* para-polymorphisms across the CLAG3 gene and HbS genotype, following meta-analysis across four datasets. Results were computed by multinomial logistic regression within each dataset using STAN, using two parasite genotype outcome levels (para-homozygous alternate and para-heterozygous, para-homozygous reference as a baseline) and using HbS genotype as the predictor. As above we assumed a dominant model of association i.e. grouping HbAS and HbSS genotypes. No additional covariates were included. The combined-evidence (log10 P-value, y-axis) was computed by meta-analysis across datasets at each *P. falciparum* variant. The positions of exons within the CLAG3 gene are shown along the bottom of the plot. **D**. The positions of predicted protein domains in the CLAG3 protein shown in grey. The first row indicates domains as predicted by Interpro, showing signal peptide (SP), non-cytoplasmic, transmembrane (TM) and cytoplasmic domains. The second row shows results of a published structural analysis of CLAG3 in its native form in complex with RhopH2 and RhopH3. Sites of strong association with HbS are linked between A and D using arrows. A known hypervariable region (HVR), exposed on the erythrocyte surface, is shown in blue. **E**. relative strength of evidence of models comparing modes of effect between para-heterozygous and para-homozygous genotypes at the chr3:124,242/140,167 site. These included: similar effects (‘dominant’), an intermediate effect in para-heterozygotes (‘dosage’), independent effects (‘independent’) and an effect restricted to each genotype (‘para-het only’ and ‘para-hom only’).

To better understand the context of this association signal, we generated long-read assemblies for two samples in the GAMCC resource and, together with available long-read genome assemblies^18^, created a multiple sequence alignment of encoded CLAG3 protein sequences (**Fig. S10 & Methods)**. An immediately notable feature is that the lead HbS-associated polymorphism in *CLAG3*.*1* (chr3:140,167 T>G; I1254R in the amino acid sequence), like many other mutations across this gene, is polymorphic in the paralogous gene copy *CLAG3*.*2* as well (at chr3:124,242 T>G / I1253R; **Fig. S10**). We refer to variants of this type, i.e. loci which show the same polymorphic alleles at paralogous locations in both genes, as ‘para-polymorphisms’. Although the presence of para-polymorphism is initially surprising, similar patterns have been observed at other paralogous gene copies in *P. falciparum*^19-21^ and, in the case of *CLAG3*, might arise through gene conversion or be transferred between genes by recombination involving *CLAG3h* hybrids. The CLAG3 proteins contained 49 para-polymorphisms and we found no fixed amino-acid differences between *CLAG3*.*1* and *CLAG3*.*2*. Among these assemblies, the lead HbS-associated polymorphism did not appear in *CLAG3h* samples, likely due to the limited number of assembled isolates used (**Fig. S10)**.

The genetic context of *CLAG3* is therefore complex, so we sought to re-analyse the HbS-association considering both paralogy and the presence of structural variation. We first used a targeted strategy to call para-polymorphic genetic variants across the samples in our meta-analysis. This approach works by re-aligning all local reads separately to the *CLAG3*.*1* and *CLAG3*.*2* sequences (as taken from the Pf3D7 reference), merging locations across the gene and then calling genotypes assuming a diploid model. This generates a new set of genotype calls across the combined *CLAG3* gene with the property that samples with different alleles at para-polymorphic loci have heterozygous genotypes in the output. We note this method does not distinguish which gene copy carries which allele.

We also assessed the presence of the *CLAG3h* deletion variant by comparing sequencing depth within the deletion region to that in flanking non-structurally varying regions. We found we could effectively identify samples carrying a single *CLAG3* copy by the presence of a low deletion sequencing depth ratio and low heterozygosity (**Fig. S11A**). The frequency of the CLAG3h deletion varied substantially between populations, with the highest frequency of around 40% in Ghana (**Fig. S11B)**. Notably, the frequency of CLAG3h was lower in the CP1 Gambian samples (16%) compared to the GAMCC (23%) which were collected at an earlier time point, an opposing trend to that seen for the *Pfsa+* mutations in The Gambia over time (**Fig. S7A)**.

We then used these calls to re-assess the association with HbS. We observed that the association does not appear to be driven by the *CLAG3h* deletion (*P*=0.037) (**Fig. 5B)**. In contrast, using the para-polymorphic genotype calls substantially strengthened the association signal at the same lead variant identified above (*P*=3.8×10^−17^, based on *N*=5,343 unmixed samples (*F*_$%_ calculated locally to the *CLAG3* region > 0.5), computed using multinomial logistic regression model implemented in STAN with -/- baseline and +/- and +/+ as outcomes; **Fig. 5B** and **Supplementary Table S3B**). Both +/- and +/+ para-genotypes showed similar effects relative to -/- (**Figs. 5B & Supplementary Table S3**; posterior probability of a dominant effect = 0.85; **Fig. 5E**), leading to a further strengthening of signal when these effects are combined (*P*=2.4×10^−19^). Although the two *CLAG3* genes copies are thought to undergo mutually exclusive expression, these results suggest the HbS-association is similar whether one or both genes carry the HbS-associated mutation. One possibility is that parasites preferentially express the CLAG3 copy carrying *Pfsa*+ when available, but detailed transcriptional data in the context of HbS infections will be required to resolve this. A second strong association was also noted at chr3:139,653 (*CLAG3*.*1*) / chr3:123,737 (*CLAG3*.*2*) C>A (*P*=4.3×10^−14^), which was also a para-polymorphic site (**Fig. S10**).

### HbS-associated mutations in the *FIKK* family

In addition to *CLAG3*, our analysis implicates mutations in two *FIKK* kinase family members: *FIKK3* on chromosome 3 and, as previously reported ^5^, *FIKK4*.*2* on chromosome 4. Like *CLAGs, FIKK* genes form a family that has expanded within the *Laverania* lineage, with approximately 21 paralogous members in *P. falciparum* ^18^. Kinase activity, substrate specificity and localisation vary across the family; *FIKK3* is thought to localise to the rhoptries, while *FIKK4*.*2* localises to the red blood cell periphery. Both genes have been observed to have threonine / serine / tyrosine specificities ^22^. In both genes, the HbS-associated polymorphisms alter the amino acid sequence but lie outside the kinase domains, which raises questions as to their functional impact. The *FIKK3* mutation (chr3:79,845 G>A) encodes serine>phenylalanine (position 137) which differ greatly in size and hydropathy properties. However, this occurs in a region that is not currently confidently predicted by structure prediction tools ^23^.

The lead *Pfsa* mutation in *FIKK4*.*2* (chr4:1,121,472 T>A) encodes leucine>isoleucine at position 960, potentially a relatively minor change. However, close to this site, *FIKK4*.*2* also contains a long hexameric repeat sequence which varies in copy number between assemblies^5^. In Methods we detail a method to call this variation in the study datasets by leveraging the available genome assemblies. We found that the *Pfsa4+* allele was strongly associated with a higher copy number of this segment in all study populations (e.g. *P* < 2 × 10^−16^ for copy number > 1 in Ghana; 42 of 44 *Pfsa4+* parasites have copy number > 1; similar results in other datasets), as well as with other nearby mutations. However, the copy number variant was not itself strongly associated with HbS (P > 0.01). These analyses suggest the possibility that the *Pfsa4+* association is driven by a complex haplotype carrying a relatively long hexamer repeat segment at this locus. We caution that this variation can only be partly resolved using short-read sequencing, and long-read sequencing approaches may be needed to fully analyse this region.

## Discussion

Malaria parasites evolve in response to a wide variety of selection pressures, including environmental changes engendered by the protective effects of human genetic polymorphisms. Over time, this may drive the emergence of parasite mutations that ameliorate or overcome these protective effects. The ultimate evolutionary outcomes of this are not known, but new datasets of host and parasite genetic variation are enabling insights into this process. For *P. falciparum*, the malaria parasite which causes the highest burden of disease and death, previous work has identified a set of genomic loci collectively termed *Pfsa* which strongly associate with infections of HbS-carriers^4,5^.

Here, we build on this by generating new whole-genome sequence data and conducting a meta-analysis of HbS-*P. falciparum* genetic association across 6,289 samples from four populations across Africa. Our results implicate genetic variation in multiple additional genome regions as associated with infections of HbS individuals, with the strongest new signal found in *CLAG3* (**Fig. 2**). While the new associations have weaker marginal association effects compared to those originally identified (with *OR*=3.9-11.4 compared to *OR*=16.9-25.1), most are supported by data from multiple populations (**Fig. S4**) and, like the originally identified *Pfsa* alleles, show unusually elevated between-locus linkage disequilibrium (**Fig. S5**), pointing to a substantially more complex genetic landscape than previously suspected.

It is not clear why such a large number of mutations are associated with HbS. One possible model is that parasite HbS-adaption might involve initial strong “driver” mutations with subsequent evolution of weaker modulatory alleles. Among the newly identified mutations, the *CLAG3* variant showed the clearest impact, substantially amplifying risk of disease in HbS individuals in combination with other *Pfsa*1/3+ alleles (**Fig. 3**). However, this effect is still much weaker than the effects detected at the *Pfsa*1+ and *Pfsa*3+ loci, making these promising candidates for the primary adaptions that enable parasites to overcome HbS. Consistent with this, our analysis of combinations of *Pfsa* genotypes across datasets suggests that *Pfsa*3+ remains the only mutation with clear evidence of an impact on HbS-resistance in isolation (**Fig. S8**). However, we caution that even the combined dataset analysed here only contains N=126 individuals with HbAS/SS genotypes, and further data is needed to fully clarify how these mutations jointly impact the protective effect of HbS overtime and across populations.

The identification of HbS-associated mutations in *CLAG3* genes is particularly note-worthy given these genes play an essential role in host-cell remodelling and nutrient uptake. CLAG3 was recently the focus of high throughput small molecule screens aimed at identifying new druggable targets^24^. Notably, both HbS-associated para-polymorphisms in *CLAG3* cause amino acid changes: the lead variant (arginine-to-isoleucine at position 1254 (CLAG3.2) / 1253 (CLAG3.1)) falls downstream of a transmembrane domain (TM), and the second mutation (isoleucine-to-leucine at position 1085/1083) sits upstream of a hypervariable region (HVR) (**Fig. 5D**). During trafficking to the infected erythrocyte membrane, CLAG3 is in complex with RhopH2 (interfacing with the middle domain) and RhopH3 (interfacing with the N-terminal domain). Within CLAG3, a helical bundle structure which contains the two HbS-associated sites encapsulates the TM and HVR regions^25^ (**Fig. 5D)**. Although the precise mechanism of membrane insertion remains unresolved, structural studies suggest that conformational changes may enable the insertion of the TM into the host cell membrane and exposure of the HVR peptide on the extracellular surface^25,26^. It is therefore possible that the HbS-associated mutations influence these dynamics.

In addition to *ACS8* (at the *Pfsa*1 locus), HbS-associated variants in two *CLAG* genes and a second *FIKK* gene implicates three major *P. falciparum* multigene families in parasite adaptation to the natural protection conferred by HbS, alongside several additional putative loci. Overall, these findings suggest a complex, multigenic adaptation to a single host polymorphism, and highlight the genomic complexity underlying host–parasite co-evolution.

## Acknowledgements

We thank the participants of the GAMCC study and all contributors to other collections used in this work. This publication made use of public data released by MalariaGEN (Pf7 ^9^ and CP1 ^4^), from Hamilton *et al*. (2023)^5^, and Saelens *et al*. (2021)^11^. Computation used the Biomedical Research Computing facility of the Medical Sciences Division, University of Oxford.

## Author contributions

Conceptualisation: AJF, GB. Resources AJF, JD, KH, AT, DB, ZZ, WLH, LA-E. Methodology: AJF, GB. Formal analysis: AJF. Software: AJF, GB. Visualisation: AJF. Supervision: GB, AJM, AA-N. Writing original draft: AJF. Editing: AJF and GB in collaboration with all authors.

## Data availability

Short read sequence read data from selective whole-genome amplification and sequencing of 905 *P. falciparum* genomes and long-read sequence data from genomes in the GAMCC dataset will be deposited on the European Nucleotide Archive (study accession: PRJEB97067). HbS and HbC genotypes are available within an accompanying sample file. A dataset of joint calls made across the GAMCC and CP1 is available from Zenodo (10.5281/zenodo.17056084). Association test summary statistics for a GWAS of severity in the GAMCC (**Fig. 1**) and the meta-analysis of HbS-associations across multiple datasets (**Fig. 2**) are also available from Zenodo (10.5281/zenodo.17056194).

## Code availability

Source code for BINGWA and HPTEST is available at https://code.enkre.net/qctool. Analysis scripts and snakemake{Mölder, 2021 #76} pipelines reflecting the analysis of the CLAG3 locus will be made available via GitHub: [link-to-be-inserted].

## Supplementary Materials

Methods, Supplementary Information, Supplementary Figures and Supplementary Tables 4-7 can be found in the enclosed document “Methods and Supplementary Information.pdf”. Supplementary data is available in “Supplementary Tables 1-3.xlsx”.

