## Supplementary material for "Associations between sickle haemoglobin and the *Plasmodium falciparum CLAG* and *FIKK* gene families revealed by meta-analysis of 6,289 African samples": Methods and Supplementary Information

**Investigation of the correlates of severity in the *P. falciparum* genome** 23
Genome-wide association analysis in the GAMCC dataset 23
Replication of investigation for severity effects using overlapping Kenyan parasite genomes 23
A putative severity variant on chromosome 1 in the *P. falciparum* genome 24

**Discussion of HbS-association evidence at the *Pfsa*1-4 loci** 24

**Assessment of independence of association and linkage disequilibrium between *Pfsa* loci on chromosome 2 and 3** 25
Chromosome 3 loci 26
Chromosome 2 loci 26

***Supplementary Figures*** 28

**Figure S1 – Available datasets for analysis of HbS-associations within the *P. falciparum* genome** 28

**Figure S2 – Detail of chr1 association with severe vs. non-severe disease in the GAMCC dataset** 29

**Figure S3 – Frequentist fixed effect meta-analysis of association with HbS** 30

**Figure S4- Combined association and geographic evidence at novel putative *Pfsa* loci** 31

**Figure S5 – Positive linkage disequilibrium between known *Pfsa* loci and putative novel *Pfsa* loci in four datasets** 32

**Figure S6 - Local linkage disequilibrium (R) between nearby known and novel putative *Pfsa* variants** 33

**Figure S7 – Temporal change in allele frequency at known and novel putative *Pfsa* loci in the Gambia** 34

**Figure S8 – Estimated multiplicative effect of putative *Pfsa* loci on the *Pfsa*1/3 background.** 35

**Figure S9 – Estimated ratio of relative risk (RRR) across different *Pfsa* genotype combinations across all datasets** 36

**Figure S10 – Multiple sequence alignment of *CLAG3* gene copies** 37

**Figure S11 – Results of CLAG3h calling across datasets** 38

**Figure S12 - Example Sanger-sequencing traces used to genotype the HbS locus.** 39

**Figure S13 – Example reads aligning to the CLAG3 region** 39

**Figure S14 – Comparison of paralogy-aware and unaware genotype calls at CLAG3** 40

**Figure S15 - Estimated ratio of relative risk of malaria based on HbS for different *Pfsa* genotype combinations** 41

**Figure S16 - Power and true discovery rate for parasite genetic effects on severity** 42

**Figure S17 - Replication of HbS association evidence across the five datasets** 43

**Figure S18 - Conditional analysis of chromosome 3 *Pfsa* association signals** 44

**Figure S19 - Conditional analysis of chromosome *Pfsa* association signals** 45

***Supplementary Tables*** 46

**Supplementary Tables 1-3** 46

**Supplementary Table 4** 46

**Supplementary Table 5** 46

**Supplementary Table 6** 47

**Supplementary Table 7** 48

*References* ......................................................................................................................... *49*

### Methods

#### Building the GAMCC dataset

##### Overview of samples

The Gambian Malaria Cases and Controls (GAMCC) samples have been previously described [1]. In brief, malaria cases with and without severe disease symptoms (described as mild within the original publication) were collected in children up to the age of 10 from Royal Victoria Hospital (Banjul, The Gambia) and the Medical Research Council Hospital (Fajara, The Gambia) during months of high malaria transmission (August to November) in 1988-90 [1]. Severe symptoms were classified as:

i. Cerebral malaria (CM) (with an unrousable coma score < 3 by standard criteria or repeated prolonged seizures lasting > 30min).
ii. Severe malarial anaemia (haemoglobin level <5gdl-1).
iii. Hypoglycaemia (blood glucose level < 2.2mmol).

Non-severe cases were considered those with a parasitaemia > 2500/ul (a level described to be associated with illness in this setting) but did not otherwise involve any of the three attributes defining severe disease (i – iii) [1].

##### Ethical approvals

Genetic sequencing of human and parasite derived DNA preserved in the GAMCC samples was approved by the Gambia Government/MRC Joint ethics Committee in May 2022 (Reference: 26354) and the Oxford Tropical Research Ethics Committee (Oxford, UK).

##### DNA preparation and selective whole-genome amplification

GAMCC samples were extracted from venous blood draws and thus contain a large proportion of human genomic DNA. First, 5mC-methylated DNA, which is present at much higher concentrations within the human compared to the *P. falciparum* genome, was specifically digested using the endonuclease McrBC as previously described [2]. 100ng of DNA, 10units of McrBC, 1 x NEBuffer2, 0.5mL of 100xBSA and 0.5mL of 100xGTP were combined in a 30µL reaction and heated at 37°C for 120 minutes, followed by 20 minutes of enzyme inactivation at 65°C.

Next, parasite DNA was enriched within samples using selective whole genome amplification (sWGA). sWGA reactions were performed as previously described with a few minor modifications [3]. In addition to the originally published primer set 10A, 6A was also used as trialled in [3, 4]. To a final volume of 50uL an sWGA master mix was added to the 50ng of McrBC reaction digest (normalised using nuclease free water) with the concentrations of 1 x NEB Phi29 buffer, 0.1mg/mL bovine serum albumin or recombinant albumin, 2.5uM of each primer pool, 1mM of dNTP mixture and 10 units of Phi29 enzyme. dNTP ratios were adjusted to 70% A/T [2].

Following this, produced mass was quantified using a PicoGreen assay following the manufacturer's instructions. Samples that did not produce an expected yield (<~40ng/µL) were re-processed. 13 samples produced no yield after two rounds of sWGA and were thus excluded.

##### Short-read sequencing *P. falciparum* genomes using Illumina

To prepare samples for sequencing, sWGA reactions were first purified using Beckman SPRIselect beads with a ratio of 1.8×. 100ng of gDNA was then used to prepare libraries. For fragmentation, the enzyme was diluted 1:2 and incubated with the sample for 8 minutes to achieve an average fragment size of 450bp. Size-selection was adjusted to recover ~550bp and eluted in 15µL. The sample was then split and 7.5µL was taken forward into a full volume PCR performed at 6 cycles also using inhouse unique dual indexing primers [5]. Post-PCR purification was performed using Beckman SPRIselect.

All libraries were then normalised using a PicoGreen or Quantifluor assay and pooled. The size profile of the pooled library was analysed on a TapeStation. The pooled library was quantified using Qubit (Invitrogen) and diluted to ~10nM for storage. The 10nM library was denatured and further diluted prior to loading on the sequencer. Paired-end sequencing was performed using a NovaSeq6000 platform (Illumina, NovaSeq 6000 SP Reagent Kit v1.5 (300 cycles)).

**Short read sequence data alignment and processing**

*P. falciparum* WGS data was processed by aligning to a concatenated assembly including the human genome with decoy HLA (GRCh38 version GCA_000001405.15), the *P. falciparum* 3D7 reference (Pf3D7 version 3) and the PhiX-174 genome (accession NC_001422). The bacteriophage PhiX genome was included because its DNA was spiked into DNA libraries in order to add complexity (PhiX has a balanced nucleotide composition), enhancing sequencing chemistry. This tended to have highly minimal coverage in our data.

The 3D7 reference is ~23Mb in length and contains 14 nuclear chromosomes as well as a mitochondrion and apicoplast genome [6]. Regions of the genome are divided into core, sub-telomeric hypervariable, sub-telomeric repeat or centromeric regions. 'Core' regions of the *P. falciparum* genome, which make up about 90% of the genome total (20.8Mb), are defined as sections of the 3D7 assembly to which short read Illumina data can be accurately aligned to, as previously annotated [7]. The analysis presented in this paper is restricted to core regions of the genome.

To process the large amounts of generated short read sequencing data in a consistent fashion we generated pipelines using the workflow tool 'snakemake' [8]. Duplicate reads were first removed from raw sequencing reads in fastq format using 'FastUniq' [9] and the adapters removed using 'Trimmomatic' [10]. 'FastQC' and 'MultiQC' [11] was used prior and post-duplicate and adapter removal to allow manual inspection of effectiveness. The cleaned reads were then aligned to the concatenated assembly (see above) using the bwa-mem algorithm [12] and the resulting files converted to a sorted BAM format using 'samtools' [13].

Certain libraries were sequenced in multiple rounds, and so data produced from multiple runs was merged following alignment. First it was checked that each bam file produced for the same sample over multiple runs came from the same source (i.e. sample IDs had not become mismatched between runs) using the tool 'BamMatcher' [14]. This calls variation within alignments (by roughly aligning to the reference sequence and calling variation using GATK) and outputs a contingency table of variation compared at each site, to give a fraction in common. Only samples which were determined to match (i.e. with "bam files are very likely from the same source" within the output report) were merged using samtools 'merge'. As well as matching, some pairs were reported to also show "the possibility of allele specific genotype" which we believe is likely due to difficulties aligning to non-core and low complexity regions of the *P. falciparum* genome.

**Long-read sequencing of two parasite genomes in the GAMCC using PacBio**

Two samples in the GAMCC which had undergone sWGA underwent long-read sequencing on the PacBio platform. The sWGA sample was first normalised to 300ng to a final volume of 50uL in nuclease free water. DNA was then fragmented to 10kB at 1500 rotations per minute for 5 minutes using the 1600 MiniG Homogenizer. Samples that had been identified as containing 1.6Kb fragments (or smaller) were input to a size selection step using Ampure PB beads (diluted to 35% volume to volume) at a ratio of 3.1x to remove fragments <5Kb. Fragment size distribution was then checked using the Agilent Femto Pulse.

Fragmented samples were prepared using the PacBio SMRTBell Prep. Kit 3.0 and indexed with the SMRTbell Barcoded Adapter Plate 3.0. Libraries were then pooled and prepared for sequencing using the Sequel II binding kit 3.2. Libraries were loaded at a concentration of 105pM with adaptive loading and were sequenced using the PacBio Sequel IIe platform with a 30-hour movie time (time of sequencing reaction).

To generate regional assemblies, we used HiFiASM v0.21.1-r686 [15] with default options to generate an assembly graph in GFA format. To appropriately order and orient the contigs in the same way as the 3D7 reference assembly, we extracted the sequences from S records and used minimap2 to align them to Pf3D7_v3. A custom script was used to then re-name and (where necessary) reverse complement sequences so that they approximately correspond to the Pf3D7_v3 nuclear, mitochondrial and apicoplast contigs; where multiple contigs aligned to the same chromosome they were given numerical suffixes. We then used liftoff [16] to lift over gene annotations from Pf3D7_v3 to the new assembly.

To analyse specific regions of interest, we first picked a focus location (usually the lead HbS-associated SNP from our analysis) and extract a 101bp kmer centred at this SNP from the Pf3D7_v3 reference sequence. We used BLAT v35 [17] to align this to all generated assemblies as well as previously published assemblies [18].

**Determination of HBB genotypes in the GAMCC samples using Sanger sequencing**

The HbS (rs334: chr11:5,248,232 T>A in GRCh37 co-ordinates) and HbC (rs33930165; chr11:5,248,233 C>T) genotypes of the majority of the GAMCC samples were determined using a PCR amplicon approach to resequence a portion of the *HBB* gene as described below. In total 701 of 903 samples were genotyped this way (summarised in **Supplementary Table 5**).

The approach used has been previously described [19, 20]. In brief, for each sample, the region was first amplified via PCR using the primers acgttggatgGTCTCCTTAAACCTGTCTTG and acgttggatgTCAAACAGACACCATGGTGC. To a final volume of 25µL, a master mix of 1.55 × BioTaq buffer, 1.24mM dNTPs, 3.1mM $MgCl_2$, 0.3uM of each primer and 0.4U/sample of BioTaq enzyme was added to a 2µL aliquot of sample (which had been pre-diluted with nuclease free water to ratio of 1:15). The PCR program is shown in **Table S1**. The original protocol was adapted to perform more reaction repeats (35 instead of 29) to generate more product

During initial testing, products were checked on a gel for a 162bp band prior to sequencing. As the project progressed and the assay became reliable, we instead used the Agilent TapeStation platform to assess a random subset of 10 samples per plate and verified that the majority had produced an amplicon. Plates were sequenced unidirectionally using the primer acgttggatgGTCTCCTTAAACCTGTCTTG by Azenta Life Sciences. On each 96 well plate a positive HbAA control (human DNA from CEPH) and a negative control (nuclease free water) was included. Sequencing was repeated once if the first attempt failed to produce usable data.

For each sample, we inspected Sanger traces using the software 4peaks, and sample genotypes were determined by inspection. An example of an HbAS trace is shown in **Figure S12A,** and an example of a reference genotype (HbAA) is shown in **Figure S12B**. Some traces showed noise across relevant parts of the amplicon and/or were supplied with QC warnings by the sequencing provider; samples were excluded from the genotyping if the genotypes could not be determined.

There were two instances of positive controls not producing any sequence, and the negative control showing an amplicon. This amplicon appears genetically identical to that of the used positive control, and it is therefore our belief that this was due to a sample mix-up between the positive and negative control on these plates. Low levels of poor-quality sequence signal were also occasionally seen within negative controls which may be suggestive of minor contamination. The low signal observed in the Sanger run indicates minimal material was present in the well, indicating that contamination would likely have occurred after the amplification step and is unlikely to have caused false positive calls that could significantly impact the analysis. Notably, no samples identified as carrying one or two HbS alleles were in adjacent wells, reducing the likelihood of cross-contamination between wells causing spurious signals.

##### Determination of HbS genotypes by whole-genome human typing

Two additional assays were applied to a subset of GAMCC samples and were used to generate HbS genotypes (**Table S5**). A subset of 122 samples with *P. falciparum* WGS data were genotyped using the ThermoFisher Axiom Precision Medicine Array, which directly assays the HbS locus. In addition, a subset of 139 samples (including the above 122) were genotyped using a low-coverage whole genome sequencing (lcWGS) approach followed by genome-wide imputation using the software QUILT [21]. We used these genotype calls for samples without accurate HbS typing from Sanger sequencing.

#### Generating a combined variant call sets to assess HbS-Pfsa interactions

##### Description of available datasets

To test for association between HbS and parasite genetic, variation we used five datasets in total:

- The GAMCC dataset described above.
- Previously published parasite sequencing and HbS genotypes from N=2,235 and N=1,936 severe malaria cases ascertained in The Gambia and Kenya in 1995-2009 [22]. HbS genotypes for these samples were determined using the Agena Biosciences MassARRAY platform. These samples were analysed as part of MalariaGEN CP1 project.
- Previously published HbS genotypes from N=1,555 mild malaria cases ascertained in Ghana in 2015-2018 [22]. HbS genotypes were determined using a Sanger sequencing approach like that described above. Parasite sequencing for these samples was published as part of MalariaGEN Pf7 [23].
- Previously published RNA-seq data from 32 infections of Malian children with mild disease, ascertained in 2016-2018 under a clinical trial (NCT02645604), including 16 individuals with HbAS genotype and 16 with HbAA genotype. These samples underwent poly-T RNA capture and sequencing at 150bp PE on the Illumina Novaseq platform [24].

For the Ghanaian dataset, the variant calls released as part of the MalariaGEN Pf7 resource [23] were used for analysis. For the GAMCC, CP1 and Malian datasets, variant calls were generated as described below. Used workflows are also available as snakemake pipeline files on GitHub as described in the **Data Availability** section.

##### Calling variation jointly in the GAMCC and CP1 datasets

To maximise variant discovery and calling accuracy in the GAMCC and CP1 sets, we implemented a joint genotype calling pipeline across these samples. In total 5,074 samples were included. In brief, we downloaded FASTQ files from samples in the published "analysis" set of CP1 and processed them together with those for GAMCC using a workflow like that used for the Pf7 dataset [23]. All reads were aligned to a combined Pf3D7_v3 (*P.falciparum*) and GRCh38 (human) genome using 'bwa mem' as described in the 'Short read sequence data alignment anr processing' section. We then applied GATK base quality score recalibration (BQSR) over each chromosome for each sequencing run for each sample. We used GATK HaplotypeCaller to call genotypes per sample and samples, before grouping and calling variation across the datasets using the GenotypeGVCF. Although *P. falciparum* parasites exist as haploid during the human portion of infection, they can also contain mixtures due to co- or superinfection, and we followed standard practice [23] by calling parasites as if they were diploid. This provides a basic way to account for levels of mixture within malaria parasite infections by inspecting heterozygous genotype calls. We then annotated variants by their region type (core, subtelomeric hypervariable, subtelomeric repeat or centromeric regions [7]), and their functional impact using 'SNPEFF' [25]. To accelerate processing this was performed in 200Kb segments of genome and the resulting VCF files were merged into chromosomes using 'bcftools'.

To create a curated call set, we used GATK variant quality score recalibration (VQSR) [26] to score genetic variants based on multiple metrics including quality normalised by depth (QD), strand odds ratio (SOR) and fisher strand (FS) measures. A set of high confidence calls published from crosses of laboratory strains (3D7 with HB3, 7G8 with GB4 and HB3 with Dd2) [7] was used as a true variant set for this analysis. VQSR was performed for separately for SNPs and INDELs separately for each chromosome in the *P. falciparum* genome. VQSR 'PASS' corresponds to retaining variants with a VQSLOD above the 99.0 tranche cutoff determined from the model.

The joint analysis callset contains 5,074 samples and may be useful for future studies. It is available as detailed in **Data Availability**.

##### Calling variation in RNA-seq data from Mali

Sequence reads from the Mali RNA-seq data were first aligned to the reference assemblies using STAR v2.7.10b [27] and the following options: –chimSegmentMin 20, --outSAMStrandField, --alignIntronMin 5, --alignIntronMax 1200, --outFilterScoreMinOverLread 0, -outFilterMatchNminOverLread 0, --outFilterMatchNmin 30, and --outFilterMismatchNmax 10 following the original analysis [24]. For this purpose we combined the human GRCh38 and parasite Pf3D7_v3 reference sequences, and the transcript annotation from GENCODE [28] v45 and PlasmoDB [29] v65, and used these as input to the STAR method.

Variants in the RNA-seq dataset were then called from the BAM files using the bcftools 'mpileup' and 'call' commands. Only reads with a mapping quality of 30 or more and bases with a base quality of at least 20 were included. INDELs were not called as part of this pipeline. Because this data was generated from RNA-seq data, certain regions were poorly covered due to variation in expression. To help ensure this did not lead to unreliable variant calls, sites were filtered so that calls with less than 5 reads ($DP < 5$) or an allele depth ratio less than 4/5 (i.e. 4 out of 5 reads are not concordant for that allele call, $AD/DP < 0.8$) were set to missing.

##### Sample filtering

Pre- and post-filtering sample sizes for each dataset are given in **Supplementary Table 1.** For samples in the MalariaGEN CP1 and Pf7 sets, the previously published "analysis" and "QC pass" sets were relied upon for sample selection in this analysis [22, 23]. CP1 samples were excluded by the following criteria:

i. Samples where the GATK v.3.8.0 CallableLoci metric (defined as the proportion of genomic bases in the core regions of the *P. falciparum* genome with at least 5× fold coverage and where at least 90% of covering reads have a mapping quality score ≥ 10) was less than 50% were removed.
ii. Samples which had >5% genotype missingness were excluded.

In addition to criteria (i), Pf7 samples which did not have a high proportion of singleton variant calls were retained in the QC pass set.

Once GAMCC samples were sequenced, they were filtered by the criteria (i) and (ii) with one change; for the GAMCC samples which had genotype missingness >10% were excluded. This more lenient threshold was adopted to retain as many samples as possible given the relatively small size of the GAMCC set. Furthermore, the potential DNA degradation due to the age of the samples may have caused dropout in certain regions without necessarily compromising accuracy at other loci. Moreover, the CP1 samples were sequenced to very high coverage (with many exceeding 100 × fold coverage, compared to on average 55 ± 18.4 × fold coverage for the GAMCC samples), which may lead to a higher proportion of missing data.

All samples from the Malian dataset were included, but strict variant calling (described below) was performed to ensure variant calls made using RNA-seq likely reflect true sample genotypes.

##### Variant filtering

For genome-wide analysis described below, we further filtered the combined callset to remove potentially spurious genotypes. First, in the combined GAMCC and CP1 call set, we used the 'vcffilter' command from the vcflib library to remove alleles with an allele count of less than 3 (AC < 3) and used the 'vt' software command 'decompose' to split multiallelic calls into separate records. This step has the effect of rescuing largely biallelic sites which were called as multiallelic due to presence of an observed rare allele (which in some cases may be the artefact of sequencing error).

Similar to previous observations [22], this dataset contains a high number of INDELs many of which are likely spurious artifacts of read misalignments in high A/T content regions. These indels also affect nearby variants often causing apparent multi-allelic variants at nearby locations. A conservative approach is to exclude these variants, and for our main genome-wide analysis we therefore restricted our analysis to bi-allelic SNPs.

These callsets also contain heterozygous calls which reflect possible mixed infections. We excluded these genotype calls from association analysis. In addition, variants were excluded if >10% of samples had missing genotypes after removing mixed calls. The variant calls from the RNA-seq data from Mali were filtered as described above.

#### Testing for associations between *P. falciparum* variation and severity

##### Association testing using SNPTEST

We used SNPTEST [30] to test for associations between variation in the parasite genome and severe vs. non-severe disease in the GAMCC dataset. This performs logistic regression using the genotype at each variant in the parasite genome as the predictor variable and severe or non-severe disease status as the binary outcome. It outputs an estimated effect (in the form of a log odds ratio ($\beta$)), associated standard errors and a *P*-value, depicted in **Figure 1G** in the main text. We restricted the analysis to the set of high-confidence biallelic SNP variants (VQSR 'PASS', sample missingness < 10%) with a moderate allele frequency (>7.5%) filtered from the analysable regions of the "core" *P. falciparum* genome. No additional covariates were included.

##### Replication analysis using the CP1 and Pf7 data

We attempted to replicate signals observed in the GAMCC using data from severe (MalareiaGEN CP1) and non-severe (MalariaGEN Pf7) infections from Kilifi, Kenya. These have the advantage of having been collected at the same location and (subject to sample filtering) at overlapping timepoints (collected in 1995-98 and 2005-09, $N^{CP1} = 894$, $N^{Pf7} = 445$, **Fig. 1B**) . We used genotypes from the joint calling described above for severe cases, and genotype calls from MalariaGEN Pf7 for the non-severe cases and formed an overlappiong. Because these samples were not collected in a matched design, we considered potential confounders (described in **Supplementary Text**) and ultimately used model incorporating drug resistance loci genotypes (*CRT* (chr7:403,625 K76T), *MDR1* (chr5:958,145 N86Y, chr5:958,440 Y184F and chr5:961,625 D124Y) and *DHPS-PPPK* (chr8:549,685 A437G)) as covariates. A genome-wide of severe versus within this Kenyan set suggested slight deflation of P-values (median $\lambda = 0.934$).

The MalariaGEN CP1 dataset also contains samples from Gambia, but the temporal overlap between the CP1 and Pf7 collections in Gambia is minimal **(Fig. 1A)**. An initial investigation suggests that confounding by time of collection and other factors is difficult to address, and we did not attempt to use these samples for replication analysis. See **Supplementary Information** for further details and power calculations.

#### Meta-analysis of HbS associations across populations

##### Association testing using HPTEST

We used HPTEST [22] to test each *P. falciparum* genetic variant for association with HbS, working separately within each of the five component datasets. In our main analysis we grouped HbAS and SS genotypes and used these as a predictor variable. HPTEST then tests for association between HbAS/SS and parasite genotype separately at each parasite genetic variant in a logistic regression framework. Because the meta-analysis framework used below implements a prior, we used only a very weak (LogF(0.1, 0.1)) prior on the main effect in the HPTEST analysis. We restricted testing to biallelic SNP variation in the core genome with least a 1% allele frequency and less than 25% sample missingness across the combined resource. No covariates were included in these computations. Analysis was restricted to VQSR 'PASS' samples within all datasets excluding the RNA-seq dataset from Mali which did not undergo VQSR.

##### Cross-population meta-analysis using BINGWA

To conduct meta-analysis, we updated the software BINGWA, which has been previously employed to assess the human associates of malaria severity [30], to work with HPTEST output files. BINGWA implements a Bayesian meta-analysis framework which can be used to assess both homogeneous and heterogeneous models of association, encoded using multivariate Gaussian priors, as well as a traditional fixed-effect meta-analysis. For Bayesian analyses, BINGWA specifies priors on the true effect sizes using a standard deviation parameter ($\sigma$) which we set to 2, or to 0.001 when modelling 'no effect'; and a between-dataset correlation parameter ($\rho$) which we set as described below.

To capture possible population variation in effects we evaluated nine different models of host-parasite effects. These included: (i) a 'similar effects' model assuming consistent effects across populations ($\rho$=0.99); (ii) an 'independent effects' model allowing uncorrelated effects between populations ($\rho$=0); (iii) a 'west-supported effects' and (iv) 'east-supported effects' model where evidence of effect ($\sigma = 2$) is supported by only West African (Gambia, Ghana or Mali) or East African (Kenya) populations; and five 'cohort-specific effects' models (v)-(ix) where the true effect is assumed to be restricted to each of the five tested datasets.

For each variant, BINGWA outputs a meta-analysis estimate of the effect size, standard error and P-value (computed under a frequentist fixed-effect model), and for each of the above model a Bayes Factor reflecting the relative evidence in the data for the model compared to the model of no association. A model-averaged Bayes factor ($BF_{\text{avg}}$ is also computed; we weighted models equally in this analysis. This value is displayed on **Figure 2** in the main text. For comparison, a standard frequentist fixed effects meta-analysis performed across the same data is shown in **Figure S4.**

In Figures **S4, S17, S18** and **S19** where effect estimates within individual datasets are shown, we applied the default prior of LogF(2,2) as described in the original study [22]. This prior provides mild regularization of effects, providing more stable estimates when host or parasite allele frequency are low, while still allowing for relatively large effect sizes. This estimate can therefore be more interpretable when comparing effects between populations. This prior was not used as input to the meta-analysis however to avoid any potential for bias due to the regularisation. This distinction will be indicated in the relevant figure legends.

##### Interpretation of Bayes Factors

HPTEST and BINGWA output a Bayes Factor ($BF$) as well as a P-value for each variant. The $BF$ is formally interpreted as the ratio of the probability of the observed parasite genotype data if the variant is truly associated ($A$) compared to the probability if there is no association (i.e. the effect size is zero) ($!A$):

$$BF = \frac{P(\text{data }|A)}{P(\text{data}|!\,A)}$$

Calculation of the Bayes factor requires assumptions about the distribution of true association effects; as above we use a Gaussian prior framework implemented in BINGWA to specify this.

Given the data, a natural posterior probability $P(A\ |\text{data})$ can therefore be expressed using the BF and the prior probability P(A) as follows:

$$P(A|data) = \frac{BF\ .\ P(A)}{BF\ .\ P(A) + \left(1\ -\ P(A)\right)}$$

Or, on an odds scale:

$$posterior\ odds(A|data) =\ BF\ \times\ prior\ odds(A)$$

Posterior odds can be understood as how much more likely the hypothesis is to be true compared to false and depends both on the data and on the assumed prior odds of association.

A potentially sensible prior belief might be that 1 in the approximately ~20,000 × 1 kilobase non-recombining blocks in the *P. falciparum* genome is associated to the HbS allele. Under this assumption, $BF$ =20,000 ($\log_{10}(BF)$ = 4.3) would be required to give a posterior odds of 1 (both an association and no association are equally likely or there is a posterior probability of 50%), and a value of $BF$ =1,000,000 ($\log_{10}(BF)$ = 6) would correspond to very compelling evidence that the variant is associated (with a posterior odds = 50, equivalent to a posterior probability greater than 98%). We have therefore highlighted variants with this level of evidence in **Figure 2** in the main text.

BINGWA outputs an individual $BF$ for each model. This can be interpreted as how much more likely the observed data are under that assumption about the effects, relative to no association. The average BF ($BF_{avg}$) therefore represents an overall summary of the strength of evidence for the association [30].

#### Supplementary variation calling at the *CLAG3* locus

##### Assessing coverage within the CLAG3h deletion

As part of further analysis of HbS associations at the *CLAG3* locus, we assessed levels of structural variation in the region. It has previously been identified that a *CLAG3* hybrid (CLAG3h) is carried by some parasites which deletes the intermediary genomic regions [31]. To evaluate the presence of CLAG3h within our sample sets we used bedtools 'genomecov' to compute sequence depth at every position within the deleted region (chr3:124,924-134,486 in 3D7 reference co-ordinates) and within an equal length of flanking genomic regions adjacent to the *CLAG3.1* and *CLAG3.2* genes (chr3:140,694-146,037 and chr3:114,116-119,458), which do not appear to contain structural variation based on long read assemblies (**Figure S19**) individually within each sample. The average depth within the deleted and flanking regions was then calculated and compared using the ratio of depth within the deletion to the flanking segments (referred to as 'normalised deletion coverage').

##### Paralogy-aware variant calling across the *CLAG3* genes

In addition to assessing depth, we also re-called sequence variation across the region. To do so, we extracted reads which originally mapped across the *CLAG3* locus in the reference genome from BAM/CRAM files using samtools (within ±5Kb around *CLAG3.1* and *CLAG3.2*, corresponding to chr3:114,458-145,660) and realigned them separately to each *CLAG3* copy in the 3D7 reference using the bwa mem algorithm (*CLAG3.1* was considered chr3:135,418-140,660 and *CLAG3.2* chr3:119,458-124,735). We then used bcftools ‘mpileup’ and ‘call’ to make variant calls across each *CLAG3* copy across all samples. Variant calls made for each gene copy were then intersected such that only variant calls that matched between each alignment were retained. This was performed to minimise regional mapping biases between each gene copy within the reference and to provide a more accurate interpretation of variation across the *CLAG3* gene within our data sets.

During initial attempts we found regions of *CLAG3* showed high levels of heterozygosity at variant sites which did not appear to exist within whole genome assemblies. In addition, samples that had very low coverage within the deleted region and so were likely to carry a single copy of *CLAG3* (i.e. the CLAG3h hybrid) also carried heterozygous calls at these sites. We noticed that non-reference haplotypes were carried entirely by certain reads leading to heterozygous calls (**Figure S13**). Using BLAST, we found that these reads had very high sequence similarity to regions outside of the *CLAG3* locus, including a region of chromosome 14 of the SN01 parasite (within the PfSN01_140007100 gene), chromosome 6 within the IT clone (PfIT_060036000), chromosome 4 in the GA01 assembly (PfGA01_04002990), and chromosome 7 of the 7G8 clone (Pf7G8_070006300). Additionally, a near-perfect match was observed with chromosome 10 of the HB3 clone (PfHB3_100043500).

These genes, nominally annotated as *CLAG2*, have been described in previous assemblies as a novel sixth member of the *CLAG* gene family, located in subtelomeric regions and predicted to be most closely related to the *CLAG8* gene [18]. The high sequence similarity between these reads and the novel *CLAG* clade, compared to other *CLAG* sequences, suggests that these parasites likely carry this sixth *CLAG* gene. Since this gene is not present in the 3D7 reference genome, reads derived from it were likely being erroneously mapped to the *CLAG3.1* and *CLAG3.2* regions, resulting in the observed heterozygosity. To solve this, we therefore reperformed re-alignment including a copy of sub-telomeric *CLAG* gene from Senegal (PfSN01_140007100) which appeared to remove the incorrectly mapped reads.

Within the resulting representation of genetic variation across *CLAG3,* heterozygous calls generally indicate a difference between the *CLAG3* copies (referred to in the main text as para-heterozygous), while homozygous calls reflect identical alleles across the copies (para-homozygous) or within a single copy in CLAG3h samples. This approach is analogous to variant calling on the X chromosome in the human genome, where individuals may be hemizygous (here, those carrying the *CLAG3h* deletion) or diploid (those carrying two *CLAG3* genes), possessing two copies of each genomic locus. A matrix displaying the difference between the original and renewed calls at the most sickle-associated site identified at this locus (chr3:140,167 T>G) is shown in **Figure S14**. We found that no calls that were originally classified as homozygous alternate or reference traded between these categories. Heterozygous calls did differ between the call sets, likely reflecting differences in alignment affinity between the two *CLAG3* copies.

**Calculating Fws locally to the CLAG3 region**

For further analysis of this data, we also computed a “local Fws” metric calculated within a 10Kb region either side of the *CLAG3.2* (chr3:109,097-119,097) and *CLAG3.1* (chr3:141,186-151,186) genes. We chose to do this instead of relying on published Fws values because we thought it best to re-calculate across the dataset to ensure no differences in computation lead to biases in metrics, and because clonality can vary across the parasite genome, with some regions retaining high levels of mixture even with a high Fws value (indicating the sample is broadly clonal).

For this calculation, original VCF files were re-filtered with adjusted criteria. VQSR ‘PASS’ variants with an allele count of 20 or more (AC > 19) were retained. Allele counts were then extracted from the VCF files using bcftools ‘query’, yielding counts of reads corresponding to the reference and alternate alleles at each site. For alleles with a minor allele frequency of at least 2.5% within both the combined GAMCC/CP1 dataset and the Ghanaian dataset, population heterozygosity (Hs) at each site was calculated across the entire dataset based on these counts. Then, individual sample heterozygosity (Hw) at each site was determined. The average Hw within 5% Hs bins was then plotted, and a linear regression was performed to assess the relationship between Hw and Hs. The Fws statistic was calculated as 1 minus the gradient of the resulting linear relationship [32]. Because each sample represents a subset of the population, Fws is expected to fall between 0 and 1. Within this computation, certain samples however were calculated to have Fws > 1 (indicating a sample is less heterozygous than expected). This is likely due to the small number of variant sites within the tested region leading to statistical noise but is still likely to represent a clonal sample.

**Calling CLAG3h within samples**

By plotting normalised deletion coverage (on a log scale) against proportion of heterozygous variant calls across the *CLAG3* re-alignment within each population (for samples where variant missingness across *CLAG3* < 20%), we saw that generally two clusters were formed; a cluster of very low coverage in the deletion and low levels of heterozygosity (likely to contain CLAG3h samples) and a cluster of high coverage within the deletion and higher levels of heterozygosity (likely to contain samples which carry two copies of *CLAG3*).

We therefore decided to classify samples based on *CLAG3* copy number using hierarchical density-based clustering using the python package hdbscan (minimum desired number of samples within each cluster was set to 50). As suspected, samples generally segregated into two distinct groups, however certain samples were not clustered and did not visually clearly belong in either cluster. We predicted that this may be due to mixed infections causing intermediate levels of coverage within the deletion or contributing to heterozygosity in CLAG3h samples. We therefore decided to restrict CLAG3h calling to samples where local Fws was moderate (local $F_{ws} > 0.5$) and missingness was low (variant missing proportion < 20%), which produced more tidy clustering results as shown in **Figure S14**.

**Generating a multiple sequence alignment for *CLAG3* sequences in long-read data**

Data from fifteen publicly available long-read assemblies from natural and laboratory isolates acquired from Otto *et al.* (2018) [18] and long-read data from two GAMCC samples (described above) were used to build an amino-acid multiple sequence alignment (MSA) of CLAG3 proteins. First, a 100bp region of DNA at the start and end of the annotated coding sequence was extracted from the 3D7 reference assembly *CLAG3.1* and *CLAG3.2* genes. Similar sequences were then searched across the available long read data using blast (using the blast-short task, a word size of 7 and a minimum e-value of 1). Using the outputs, the likely start and end of each gene was determined within each assembly. Because *CLAG3.1* and *CLAG3.2* share high levels of sequence identity, two strong matches were often ascertained. The first on the contig was considered to be *CLAG3.1* and the second *CLAG3.2*, matching the arrangement of the reference assembly. For some parasites, only one match would be discovered, often with high matching to the *CLAG3.1* start sequence and to the *CLAG3.2* end sequence. These isolates would be considered to be carrying the CLAG3h deletion.

Once the start and end locations of the genes in each assembly had been ascertained, the entire DNA sequence of the coding sequence was extracted. From this, a DNA MSA was generated using the program 'mafft' using the '—auto' parameters. From this alignment, the locations of the starts and ends of each exon, according to the annotation of 3D7, were extracted, and converted to amino acids using the 'SeqIO' package in python and saved in a FASTA file. Amino acid sequences were then also aligned using 'mafft' and plotted in the main text in Figure 3.

To annotate the amino acid sequences, sequences were uploaded into the InterPro online tool (https://www.ebi.ac.uk/interpro/). The hypervariable, N-terminal, middle and helical bundle regions were located based on the descriptions within Ho *et al.* (2021) [33].

#### Supplementary variation calling at the *FIKK4.2* locus

##### Assessment of complex *FIKK4.2* variation

To inspect variation at the *FIKK4.2* locus (containing the *Pfsa4+* mutation at chr4:1,121,472), we first generated a MSA across available assemblies as follows. We extracted the 101bp sequence centred at this SNP (`ACGATAATGATGACAGTGATGCAAGCGATGCAGTTCATGAAGATATTGAGT`**T**`ACTTGAGTCTTATAGTGATTTGAATAAATTTAATGAGATGTTAACAGAA`) from the Pf3D7_v3 reference assembly and aligned it to each genome assembly using BLAT. We noted that the kmer matched to chromosome 4 on all assemblies (with >= 99 matching bases) except for the PfSD01 isolate from Sudan, for which it aligned to chromosome 7 with 99 matching bases (but not to chromosome 4). This is consistent with the publicly available gene annotations for this genome, which indicate a major translocation of a large genome segment in this genome. We then expanded the alignments by adding 10kb to either side and used MAFFTv7.490 with default options to compute a MSA.

Using this MSA, we identified a set of DNA kmers which are shared identically across all isolates and marked the start and end of the coding sequence of *FIKK4.2* in each exon (detailed in the table below).

| Exon | 5' kmer (DNA/*AA sequence*) | 3' kmer |
|---|---|---|
| 1 | ATGAATTATTTTTCTAAATACAAAGTTATT<br>*M N Y F S K Y K V I* | TATTTTTTGTTTATAATTCCATTG<br>*Y F L F I I P L* |
| 2 | AATGAAGTAATATACAATAAATAT<br>*N Q V I Y N K Y* | GGTCTCAAAGATATAATTAAC<br>*G L K D I I N* |
| 3 | AAATTATTAAGCCTAGAAAGT<br>*K L L S L Q S* | GAACATCCATGGTGGATTAATGAAGATTAA<br>Q H P W W I N Q D * |

Inspection of the assemblies shows that The FIKK4.2 amino acid sequence contains a long hexamer repeat segment with the majority of repeats being of the form (SD[H|N|S]NH[K|M]). The flanking and repeat sequences are identified in the table below along with Pf3D7 coordinates.

| Repeat end | DNA and AA sequence identifying FIKK4.2 Srepeat With Pf3D7_v3 coordinates **(Repeat sequence in bold)** |
|---|---|
| 5’ | 1,119,738<br>\|<br>GAAGAGGATAAGAATATGA**TGGAAAATAATCATAAG...**<br>*E E D K N M* ***S E N N H K ...*** |
| 3’ | 1,121,393<br>\|<br>**...AGTGATCATAATCACAAAAGTGATCATAAAAAAAAT**AATAACAATAATAAGGAT<br>***... S D H N H K S D H K K N*** *N N N N K D* |

Inspection of the MSA shows this repeat segment varies in copy number across the assemblies, from 402 (PfKH01) to 546 (Pf3D7) amino acids (67 - 91 hexamer repeats). Two factors contributing to this length variation are: copy number variation of a ~180bp segment which is present in variable form in three copies in Pf3D7, at approximately 1,120,534-1,120,135, but in one or two copies in other assemblies [20]. For example, the PfCD01 genome contains a single copy of this segment at PfCD01_04:1,081,668-1,081,852.

In addition to the repeat segment, we also noted a number of single nucleotide polymorphisms and indels proximal to the lead *Pfsa4+* mutation, which were excluded from the Ghana dataset in our original GWAS as they were called as multiallelic. In Pf3D7 coordinates these include SNPs at 1,121,457 C>G and 1,121,328 A>C, and an overlapping 18bp deletion variant at 1,121,311 - 1,121,328.

##### Calling *FIKK4.2* variants in short read data

Calling the *FIKK4.2* repeat variation in study samples (i.e. based on short-read sequence data and from samples that have undergone SWGA) appears challenging. To attempt this, we adopted a read alignment approach which we now outline. The approach assumes that sample genomes are structurally similar to one of the assembly genomes (across the repeat segment) and calls this variation by aligning reads to each of the assemblies. We also assessed read coverage (i.e. sequencing depth) within the copy number variable segment.

For the first approach, working in each study sample separately, we first extracted all sequence reads aligning to the *FIKK4.2* region (i.e. chr4: 1,115,000-1,125,000) from the genome-wide alignments. We then re-aligned these reads to each of the assemblies (using the same ~20kb segment used for our MSA above) and collected the resulting alignments in a single BAM file.

Using PfCD01 as a reference we then further extracted the read IDs of reads with primary alignments over the hexamer repeat segment (identified as PfCD01_4:1,080,875 – 1,082,150; after adjusting the upper end of this range to be >150bp away from the lead *Pfsa4+* SNP at PfCD01_04:1,082,325). Then, for each assembly $G$, we extracted the alignment scores of these reads from the primary alignments to the assembly and summed them to obtain a total read alignment score (denoted $AS(G)$)

of the reads to $G$. $AS(G)$ can be approximately interpreted as a log-likelihood of the read sequences given the alignment location (up to a constant), i.e. $AS(G) = \log P(\text{reads}|G)$. Therefore, assuming that one of the assemblies represents the sample genome, the probability the reads originated from genome $G$ can be obtained using Bayes' theorem:

$$P(G|\text{reads}) = \frac{P(\text{reads}|G)P(G)}{\text{normalising constant}}$$

where the normalising constant is obtained by summing the numerator over all the genomes.

To call the copy number segment, we assigned copy number of 1, 2 or 3 to each genome based on kmer sharing across the region[20] and assumed:

$$P(\text{copy number } k|\text{reads}) = \sum_{G} P(\text{copy number } k|G) \cdot P(G|\text{reads})$$

Finally, we called the sample as arising from genome $G$ or copy number $k$ if the probability computed in this way was at least 90%. Across 6,296 samples included in the GWAS and in this analysis, 6,166 were assigned a genome of origin $G$ and all but 27 were assigned a copy number call (see **Data availability**). Copy numbers were called as follows:

| Dataset | Copy number 1 | Copy number 2 | Copy number 3 | No copy number call |
|---|---|---|---|---|
| Gambia GAMCC | 675 (81%) | 142 (17%) | 14 (2%) | 0 |
| CP1 Gambia | 1505 (67%) | 691 (31%) | 34 (2%) | 5 |
| CP1 Kenya | 1788 (92%) | 138 (7%) | 8 (<1%) | 1 |
| Pf7 Ghana | 1155 (89%) | 114 (9%) | 5 (2%) | 21 |

We also compared the above calls to read coverage, based on reads aligned to PfCD01genome in the CNV segment PfCD01_04:1,081,665-1,081,835, as well as a nearby non-copy-variable segment (referred to as a 'control segment' below) at PfCD01_04:1,081,600 – 1,08,200. In principle, if the above copy number calling is accurate, we would expect to see higher coverage for higher copy numbers. We did observe this on average, with mean CNV:control segment ratio being 1.01, 1.59 and 1.63 for samples called with cn=1, cn=2, and cn=3 respectively. However, the three distributions were not distinct indicating substantial noise in this approach. This could indicate remaining inaccuracies in the above calls but may also be due to the use of selective whole genome amplification (which tends to cause high variation in coverage across the genome). On balance our interpretation is that the above approach likely captures some of the variation in repeat length, as it is represented in the assemblies, but is unlikely to be a fully accurate call of the copy number variation. In particular we caution that this analysis makes the assumption that each sample genome is similar (including structurally similar) to one or more of the assembly genomes across the repeat unit. The number of genome assemblies available is limited, and it is plausible that additional variation exists; some samples are therefore likely to be inaccurately captured by the above call. Long-read methods may be needed to fully address this region.

To capture other variants nearby the lead *Pfsa4+* mutation, we re-processed the sample VCF files in the region and used vcflib's 'vcffilter' tool to remove rare alleles (with count < 5), and bcftools norm to split remaining multi-allelic variants into biallelic records. Calling is likely unreliable within the repeat unit, so we focussed on variants at position chr4:1,120,135 or above on the 3D7 reference genome.

**Pfsa4+ and copy number linkage**

To compare linkage between the *Pfsa4+* mutation and the copy number variant, we tabulated the two variants in each dataset and used Fisher's exact test to compute a P-value.

**Testing *FIKK4.2* variants for association**

We re-tested for association between HbS genotypes and each of the above variants, as well as the *Pfsa4+* lead variant identified in the GWAS using a custom R script. In Ghana, four variants (the *Pfsa4+* lead SNP chr4:1,121,472 T>A, two other nearby SNPs at chr4:1,121,328:A>C and chr4:1,121,457:C>G and copy number > 1) were associated with HbAS/SS genotypes with $P < 1 \times 10^{-3}$. However, the *Pfsa4+* lead SNP had the strongest evidence ($P = 5 \times 10^{-10}$), and conditioning on the other variants did not remove signal at *Pfsa4+*.

This analysis also did not strongly alter the results in the other datasets: no variant was strongly associated with HbAS/SS genotypes in CP1 Kenya, CP1 Gambia or the GAMCC dataset. A very modest association was noted between copy number > 1 and HbS in the two Gambian datasets (*OR* = 1.02 and 1.03 in GAMCC and CP1 Gambia; *P* = 0.08 and 0.02 respectively).

#### Statistical analyses

**Estimating the variation in relative risk conferred by HbS across combined parasite genotypes**

Given the large number of *Pfsa+* alleles now identified, we set out to estimate the joint impact of the alleles, including those within the *FIKK3* and *CLAG3* genes, on the Although it is possible to estimate the relative risk (RR) conferred by HbS genotypes against specific parasite genotypes (under simplifying assumptions (Band *et al.* (2022)), this requires data on HbS frequencies in population control samples which were not available for the GAMCC and Ghanaian collections. We therefore instead aim to estimate ratios of these relative risks (*RRR*), thereby capturing variation in the *RR* between parasite genotype levels. Concretely, let $G$ denote the combined parasite genotype of an infection across a chosen set of loci. We will work relative to a chosen baseline genotype (e.g. the genotype carrying all the *Pfsa-* alleles at *Pfsa* loci) which we denote $G = 0$. Let $H = S$ and $H = A$ denote hosts carrying the HbS allele or not respectively. For each combined *Pfsa* genotype $G = x$ we consider the ratio

$$RRR(G = x) = \frac{P(G = x|h = S)}{P(G = x|h = A)} \Big/ \frac{P(G = 0|h = S)}{P(G = 0|h = A)} \quad \text{(E1)}$$

The two terms in the ratio are, respectively, the $RR$ conferred by HbS against disease with parasite genotype $x$ or baseline genotype 0. If the protection due to HbS is similar between genotypes, we would therefore expect to have $RRR = 1$, while (assuming no confounding effects) $RRR < 1$ or $> 1$ would indicate that the protective effect against genotype $x$ is stronger than or weaker than that for the baseline genotype.

To estimate the *RRR*, we work on the log-*RRR* scale and use a multinomial logistic framework, as we now describe. Suppose the possible combined genotypes across the chosen loci are enumerated as $g = 0,1, \cdots T - 1$, where T is the total number of possible genotypes. (In practice, we typically focus only on those genotypes that are observed in data so that T may be smaller than $2^{\#loci}$). Let f be the multivariate logistic function that maps $T$ log-odds values into $T$ probabilities:

$$f_i(q) = f_i(q_0, \cdots, q_{T-1}) = \frac{e^{q_i}}{\sum_j e^{q_j}}$$

where $q = (q_0, q_1, \cdots, q_{T-1})$ is a vector of real numbers. Note that these probabilities sum to one i.e. $\sum_{j=0}^{T} f_j\,(q) \equiv 1$, and correspondingly, the values of $f$ are unchanged if we add any constant to the entries of $q$.

For an infected individual with host genotype H and fixed covariates Z of dimension $d$, we model the frequency of combined genotype $G = x$ in terms of its log-odds as

$$P(G = x|H, Z) = f_x(\mu + H\omega + Z\gamma) \qquad \text{(E2)}$$

Here:

- $\mu = (\mu_0, \cdots, \mu_T)$ is a row vector of baseline parameters influencing the frequency of outcome genotypes for individuals with HbAA genotype.
- $\omega = (\omega_0, \cdots, \omega_T)$ is a row vector of "effect" parameters reflecting the additional log-odds of each outcome genotype in hosts with HbAS/SS genotypes
- $\gamma = (\gamma_{ij})$ is a $d \times L$ matrix of fixed effect parameters, with one row per entry of the covariates vector $Z$.

Since the probability values are unchanged after adding a constant to the parameters, we assume that $\mu_0 = \omega_0 = \gamma_{0,\cdots} \equiv 0$.

The relationship between (E1) and (E2) is as follows. For two individuals with the same covariates but with H=A and H=S respectively, it follows that

$$\frac{P(G = x|H = S, Z)}{P(G = x|H = A, Z)} = \frac{\left(\frac{\mathrm{e}^{\mu_x+\omega_x}}{\sum_y \mathrm{e}^{\mu_y+\omega_y}}\right)}{\left(\frac{\mathrm{e}^{\mu_x}}{\sum_y \mathrm{e}^{\mu_y}}\right)} = \frac{e^{\omega_x}}{\left(\frac{\sum_y \mathrm{e}^{\mu_y}}{\sum_y \mathrm{e}^{\mu_y+\omega_y}}\right)}$$

Since the denominator is the same across all genotype levels,

$$RRR(G = x) = \frac{e^{\omega_x}}{e^{\omega_0}} = e^{\omega_x}$$

as we have assumed that $\omega_0 \equiv 1$. Thus $\omega_x$ represents the log-ratio of relative risks corresponding to genotype $x$ relative to the baseline genotype. For a single bi-allelic locus, the above is equivalent to the logistic regression approach used for our GWAS.

In practice we use this framework in two ways. First, for a given set of *Pfsa* loci we use multinomial logistic regression to estimate $\omega_x$ for each combined genotype. To prevent overfitting of the multinomial model, we fit the model under a mild regularising prior (as described in [22]); specifically we assumed a multivariate Gaussian prior on the effects with a mean of 0 and a standard deviation of 10, and the correlation between parameters across the model set to 0. This prior has the effect of regularising estimates for genotypes that have low sample counts. We caution that for these genotype combinations, the resulting parameter values can be interpreted as plausible estimates given the current data, but larger samples would be needed to fully specify these effects. To implement this model, we used the TMB framework in R [34]. Results are shown in **Figure 3** (for Pfsa1-4 and *CLAG3.1*) and **Figure S9**.

As part of this research, we recalled variation at the *CLAG3* locus to account for high levels of sequence paralogy, which lead to differences in genotype classifications (**Figure S14**). To assess whether this altered our estimates of *RRR*, we repeated the analysis using the new *CLAG3* calls (restricted to homozygous genotypes) and found this to give very similar estimates (**Figure S15**).

Secondly, we also used this framework to directly estimate the contribution of specific loci, taking the full set of loci into account. Concretely, suppose we divide the loci into two subsets A and B, where we want to estimate the effect of loci in B while accounting for loci in A. For any combined genotype $x$ we let $a(x)$ and $\mathrm{b}(x)$ be the component of $x$ at loci in A and B respectively. We then assume that these two subsets contribute additively to the log *RRR* parameter:

$$\omega_x = \alpha_{a(x)} + \beta_{b(x)} \tag{E3}$$

The key aspect of this formula is that $\beta_{b(x)}$ is shared across all genotype levels $x$ that carry the same genotypes at the loci of interest B. Thus, $\beta_{b(x)}$ represents the additional log *RRR* for genotype $b(x)$ at the loci in B, compared to genotypes that are the same at A loci but have baseline genotypes at the loci in B.

A concrete example used in the main text is to take A = *Pfsa1-4* and B = *CLAG3.1*, so that we are aiming to estimate the contribution of the *CLAG3.1 Pfsa+* allele to the *RRR*. In this example, if $x = +,-,+,-,+$ across the five loci, we will have $a(x) = +,-,+,-$ and $b(x) = +$.

This formulation is different to analysing the loci in B separately from those in A, and is instead more akin to conditioning on the A loci when estimating the parameters for B. An extreme example is to consider the hypothetical situation where A and B each contain one locus which are perfectly correlated (i.e. in perfect LD), so that in effect, $a(x) = b(x)$ in equation (E3). This is analogous to the situation of perfectly correlated predictors in standard multivariate regression problems and leads to a redundancy in parameterisation of (E3).

In practice, we fit this model using a similar prior formulation as described above for the $\alpha$ parameters, and a flat (improper) flat prior on $\beta$.

**A paralogy-aware approach to association testing at the *CLAG3* locus**

In order to test for an association between carriage of the CLAG3h deletion and HbS, the 'statsmodel' package within python was used to conduct a logistic regression separately within each dataset using CLAG3h deletion status as the binary outcome and HbS genotype as the predictor using a dominant encoding (HbAS and HbSS genotypes were grouped together). Effect estimates were combined using a fixed-effect meta-analysis. The estimated effect as an odds ratio (the exponent of the computed β) and 95% confidence interval are shown in **Figure 4B** in the main text.

Association testing between HbS and parasite genotype across the *CLAG3* locus posed additional challenges. Heterozygous variant calls, which tend to represent the presence of mixed infection in *P. falciparum* genetic data, were excluded from our genome-wide assessment of HbS-associations implemented using HPTEST. However, the newly generated variant calls across the *CLAG3* locus contain para-polymorphisms (polymorphisms that differ between the paralogous *CLAG3.1* and *CLAG3.2* genes), which are encoded as heterozygous genotypes. To assess genotype associations in this data, we instead implemented a custom Bayesian multinomial logistic regression framework using Stan (implemented in Python using 'pystan'). This allowed us to estimate the effect of HbS on both para-heterozygous and para-homozygous genotypes. Genotypes at each parasite variant were therefore encoded additively (0 = para-homozygous reference, 1 = para-heterozygous, 2 = para-homozygous alternate), whereas human genotype at the HbS locus was encoded dominantly (0 = homozygous reference, 1 = heterozygous or homozygous alternate). To parse these genotypes into Python we used the package 'scikit-allel' [35].

Initially, we fitted a multinomial logistic regression model using Stan using para-homozygous reference as the baseline, producing posterior effect estimates (β) of HbS on para-heterozygotes and para-homozygote alternate genotypes ($\beta_1$, $\beta_2$). We specified weakly informative multivariate normal (MVN) priors on variance with mean 0 of variance of 100, and no enforced correlation between effects:

$$\beta \sim \mathrm{MVN}(0, \Sigma)$$

For each variant, posterior samples of β were drawn from four chains with 2,000 iterations per chain. Posterior means, variances, standard errors, and two-sided *P*-values were calculated from these samples. Variants with a minor allele frequency below 2.5% or missing genotype data in more than 15% of samples were not tested. For each retained variant, counts of samples by outcome and predictor genotype combination were reported, along with parameter estimates.

Following this initial computation, we performed a fixed effect meta-analysis using custom python code. For each variant, we extracted the β vector and the corresponding 2 × 2 covariance matrix Σ. Precision weights were calculated by inverting each population's Σ. The meta-analytic effect sizes were then calculated by multiplying the inverse of the total precision matrix (i.e., the sum of the precision matrices) by the weighted sum of effect estimates. Standard errors were computed as the square roots of the diagonal elements of the resulting meta-analytic covariance matrix. For each effect estimate ($\hat{\beta}$), a z-score was calculated as $\hat{\beta}$ divided by its standard error, and two-sided *P*-values were derived using the standard normal distribution. We then computed a joint significance over both outcome levels using the inverse of the meta-covariance matrix and comparing this to a chi-squared distribution with two degrees of freedom.

In order to interpret effect sizes, we altered the baseline of the multinomial logistic regression post-computation to reflect the carriage of the non-effect HbS-associated allele, which was sometimes the non-reference allele based on the Pf3D7 reference genome. This was implemented by subtracting the linear predictors of the new baseline from the other outcome logits, effectively rotating the parameterisation.

As well as a fixed effect meta-analysis, we also compared different models of effects between HbS and para-polymorphism. This approach was similar to that described in the above sections, used to compare models of HbS-association in the parasite genome. In contrast to the above analysis which made use of the programme BINGWA, this analysis was performed using custom python scripts. We compared six different models of effect using the follow covariance structures:

- "Independent": $\Sigma = \begin{bmatrix} 4^2 & 0 \\ 0 & 4^2 \end{bmatrix}$ (large variance, no correlation)
- "Dosage": $\Sigma = \begin{bmatrix} 2^2 & 0.99 \times 4 \times 2 \\ 0.99 \times 4 \times 2 & 4^2 \end{bmatrix}$ (para-heterozygotes show half the effect of para-homozygotes)
- "Dominant": $\Sigma = \begin{bmatrix} 4^2 & 0.99 \times 4^2 \\ 0.99 \times 4^2 & 4^2 \end{bmatrix}$ (matched effects)
- "Para-het only": $\Sigma = \begin{bmatrix} 4^2 & 0 \\ 0 & 0.01 \end{bmatrix}$ (effect restricted to para-heterozygotes)
- "Para-hom only": $\Sigma = \begin{bmatrix} 0.01 & 0 \\ 0 & 4^2 \end{bmatrix}$ (effect restricted to para-homozygotes)

For the "dosage" and "dominant" structures, we selected a correlation term of 0.99 to avoid enforcing perfect dependence between effects, allowing for minor deviations.

For each of these covariance structures, we computed the marginal likelihood of the observed β vector. A Bayes factor was computed by taking the ratio of multivariate normal densities under the posterior (data + prior) and prior-only distributions.

**Measuring genome-wide between chromosome linkage disequilibrium**

Calculating between chromosome LD iteratively between every variant pair (a total which scales quadratically with number of tested variants) in the *P. falciparum* genome would be time consuming and computationally laborious. To overcome this, we designed a series of matrix operations.

To illustrate, let $A$ and $B$ denote matrices of genotypes (as 0s, 1s and missing values) for the first and second chromosomes respectively. Matrix $A$ has the dimensions $m \times n$, where $m$ represents the number of variant sites on the first chromosome and $n$ represents the number of samples. Matrix $B$ has dimensions $p \times n$, where $p$ is number of variants on the second chromosome. Correlation is calculated using the formula:

$$Cor(A, B) = \frac{Cov(A, B)}{sd(A)\, sd(B)}$$

Covariance is calculated by:

$$Cov(A, B) = \frac{\Sigma(A - \bar{A})(B - \bar{B})}{N}$$

By our implementation, each variant (row) within $X$ and $Y$ were first mean centred and divided by their standard deviation to form matrices $X$ and $Y$ respectively. Then an unnormalised correlation is calculated by:

$$uCor = X \cdot Y^t$$

This was calculated in a single computation using the matrix operations package 'numpy' within the programming language Python. Resulting values must then be normalised by sample size ($N$ ) to produce a Pearson's correlation R value. The genetic data used in these calculations often contains missing genotypes. This can prevent computations being made, so to get around this we used masked numpy arrays which prevent missing values effecting the calculations of covariance or standard deviation. In order to get a count of samples where both genotypes were not missing, we used:

$$count = A \cdot B^t$$

The count matrix has the same dimensions as uCorr ($m \times p$). Finally, a matrix of Pearson's correlation R values for each pair was computed by: $Corr = \frac{uCor}{count}$.

#### <u>Supplementary Methods</u>

**Modelling true discovery for genetic associations and performing power analyses**

To aid in the interpretation of P-values for the GWAS of severity using the GAMCC resource presented in Figure 1 in the main text, we conducted a power analysis using simulations. Within each simulation, 2000 sets of genotypes were generated using a binomial distribution. Input population allele frequencies ($f$) and tested effect sizes ($OR$) were used to generate the allele frequency in cases ($P_{cases}$) using the formula:

$$P_{cases} = \frac{OR \times \frac{f}{1-f}}{1 + OR \times \frac{f}{1-f}}$$

This formula reflects the calculation of relative risk (RR), and how we use the OR to estimate this.

$$RR = \frac{P(severe|G = 1, S)}{P(severe|G = 0, S)}$$

Here, the RR reflects the probability of severe disease (as opposed to non-severe malaria) (denoted as 'severe' in the formula above) in infections where parasites carry a certain genotype ($G = 1$) as opposed to infections that do not ($G = 0$) in the study population '$S$'. Because severe malaria is rare, even within a population of malaria infections, the $OR$ calculated within regression analyses can be used to estimate RR. By expanding the formula above using Bayes Theorem therefore:

$$RR = \frac{P(G = 1|\ severe,S)}{P(G=0|\ severe,S)} \cdot \frac{P(G=0|S)}{P(G=1|S)} = OR$$

Therefore, because $f$ = P($G = 1|\ S$):

$$\frac{P(G = 1|\ severe, S)}{P(G = 0|\ severe, S)} = OR \times \frac{f}{1-f}$$

To convert this from odds that $G = 1$ to probability, we can use $x \rightarrow \frac{x}{1+x}$. This gives the formula for $P_{cases}$ above.

For cases, genotypes ($G$) were therefore drawn from the distribution:

$$G \sim Binomial(1, P_{cases})$$

Whereas for controls, genotypes were drawn from:

$$G \sim Binomial(1, f)$$

The number of cases and controls drawn were chosen to reflect the sample sizes of the assessed association.

Using the generated sets of genotype calls, a logistic regression was then performed, and the power and true discovery 'rate' (that is, the probability that an association is true given the p-value of the association is below a threshold) of the association assessed. For a p-value threshold denoted $T$, given that an association is true (represented as $A$), the power of a test of association can be expressed as $P(p < T\ |A)$.

By inputting this into Bayes Theorem, the probability of true discovery (given the p-value is smaller than T) can be simplified to give:

$$P(A|p < T) = \frac{power \times P(A)}{power \times P(A) + T \times \left(1 - P(A)\right)}$$

Here, P($A$) therefore refers to the prior belief in $A$, which can have a large impact on the estimated true discovery. Ideally, this prior could be learned from other studies, however this is not possible for the presented discovery analyses. We therefore estimated that 1 in approximately 20,000 non-recombining blocks in the *P. falciparum* genome might be truly associated with disease severity was adopted. Therefore, for a range of $T$ thresholds, by counting the proportion of logistic regression association results within the 2000 sets of simulated genotypes sets where p < $T$, the power was determined and using this $P(A|p < T)$ was calculated using the above formula.

### Supplementary Information

#### Investigation of the correlates of severity in the *P. falciparum* genome

##### Genome-wide association analysis in the GAMCC dataset

The GAMCC analysis set contains substantial numbers of infections with severe (N=502) and non-severe (N=329) symptoms which were classified at the time of collection (see **Methods**). By comparing the newly sequenced parasite genomes between infections from each of these categories using genome-wide association study (GWAS), it may be possible to identify parasite loci that predispose infections to increased severity. To aid in the interpretation of the results of GWAS using this data, and to help determine an appropriate *P*-value threshold to dictate whether an association might be of interest, we first conducted a power analysis. To do so we simulated sets of genotypes under the assumption of a true association, using the sample sizes available in the GAMCC, across multiple scenarios. These scenarios included two effect sizes (odds ratio (OR) = 2 and OR = 4) and varying baseline allele frequencies (5%, 7.5%, 25%, 50%). Based on association test results computed using these simulations, we determined the probability that a P-value would fall below a given threshold when an association truly exists (the power). By using Bayes Theorem, assuming a prior belief that one region in the *P. falciparum* genome was associated with severe disease, we also calculated power to estimate the probability that an observed association is true when the P-value is below the chosen threshold (the true discovery rate (TDR)) (see **Supplementary Methods)**.

We determined that a GWAS testing for severity using this data was only well powered to find large severity increasing effects at low to moderate frequencies, (5-10% allele frequency) and could not resolve severity reducing alleles at these frequencies (**Fig. S15**). This imbalance in power between effect types is due to the uneven sample sizes between phenotype categories. At large allele frequencies however (25-50%) we are moderately powered to infer both effect types. To produce a TDR of 80% across most allele frequencies for strong severity increasing effects (*OR*=4) a *P*-value threshold at roughly the Bonferroni correction threshold appeared a suitable threshold with which to interpret this analysis (displayed as a dashed line on **Fig. S15**).

Based on the above findings, we performed GWAS across a set of high confidence biallelic SNP variants (VQSLOD score > 2, sample missingness < 10%) with a moderate allele frequency (>7.5%) filtered from the analysable regions of the "core" *P. falciparum* genome. The results of this are shown in **Figure 1G** in the main text.

No variants reached the Bonferroni level of statistical significance indicated as potentially compelling by the presented power analysis ($P<1.8\times10^{-5}$). However, a variant at position 557,240 on chromosome chr1 (T>A) in the Pf3D7 reference assembly reached levels of near significance. This variant showed evidence of a protective effect (OR=0.355 (0.214-0.589)) and an allele frequency of 9% ($P=6.09\times10^{-5}$). Based on our model of TDR, we estimate that there is modest evidence of this variant being truly associated with severity at this p-value threshold (under these parameters, power=75.6% and TDR=67.9%).

##### Replication of investigation for severity effects using overlapping Kenyan parasite genomes

Since no associations of clear statistical significance were observed, we attempted to independently replicate this analysis. There are very few other parasite genomic datasets available that categorise infections based on severity phenotypes. However, two public resources of parasite WGS data curated by MalariaGEN, specifically the Consortial Project 1 (CP1) dataset of severe cases from Gambia and Kenya (first described in [22]), and the Pf7 resource of community infections [23], offered a potential opportunity to undertake replication.

The Pf7 resource was not collected under rigorous disease phenotyping definitions which poses a challenge for this analysis. For the purposes of this analysis, Pf7 samples were all considered as "non-severe" and CP1 as "severe". However, since the Pf7 samples were collected at different times to the CP1 set, this means that the severe and non-severe cases were not only stratified by disease status but also by time. Therefore, there is the potential for confounding of GWAS comparisons using these data due to genetic drift or other modes of allele frequency shift within parasite populations between time points.

Parasite genomes from both Kenya and Gambia are represented in the Pf7. Following filtering to the published QC pass set [23] and restricting samples from Kenya to those from Kilifi (matching the CP1 collection) 602 Kenyan genome and 818 Gambian genomes were available to compare to the CP1 samples. However, initial GWAS scans using these data indicated high levels of inflation (as measured by the lambda statistic ($\lambda$)). $\lambda \gg 1$ indicates an inflated GWAS model which likely occurs through the effects of systematic bias. In both populations this appeared to be driven by strong signals of association at known drug-resistant loci. To mitigate the effects of temporal variation in drug-resistant allele frequency, we focused on a set of overlapping samples from Kenya (collected in 1995-98 and 2005-09, $N^{CP1} = 894, N^{Pf7} = 445$, **Fig. 1B**). Results from comparisons in this subset produced far less inflated $\lambda$, but an association close to the chloroquine resistance locus still appeared within the results. If real, confidently untangling an association at this locus from confounding due to the spread of anti-malarial drug resistance is challenging. For our final replication analysis we therefore adapted a model incorporating drug resistance loci genotypes (*CRT* (chr7:403,625 K76T), *MDR1* (chr5:958,145 N86Y, chr5:958,440 Y184F and chr5:961,625 D124Y) and *DHPS-PPPK* (chr8:549,685 A437G)) as covariates within the restricted years set, which produced a slightly deflated model overall ($\lambda = 0.934$). GWAS was conducted using SNPTEST as described in **Methods**. For the Gambian population, temporal overlap between the CP1 and Pf7 collections is minimal **(Fig. 1A)** and similar attempts were not found to reduce $\lambda$ to a value near 1.

##### A putative severity variant on chromosome 1 in the *P. falciparum* genome

By comparing the results of both GWAS scans, the variant chr1:557,240 T>A discovered in the GAMCC set also showed evidence of association with severity within the CP1/Pf7 GWAS (**Fig. 1**C). This consistent pattern across datasets suggests a potentially genuine link between this variant and malaria severity. The variant is at the distal end of chromosome 1, falling 18.6Kb within the designated "core" region [36]. Reads in the GAMCC cohort mapping to chr1:557,240 did so with high mapping quality scores, indicating it has been confidently called and so is unlikely to be an artefact of sequencing error. It is a non-coding single nucleotide variant 459bp downstream of the protein coding gene PF3D7_0114500 of unknown function which is listed as a *Plasmodium* exported protein within hypothetical gene family 10 (*hyp10*) (**Supplementary Figure 2**) [37]. The variant is also 1802bp upstream of a VAR pseudogene (PF3D7_0114400, PfEMP1 exon 2). Protein-coding VAR genes encode PfEMP1 proteins which are believed to be significant virulence factors because they facilitate the cytoadherence of infected red blood cells to host tissues [38].

#### Discussion of HbS-association evidence at the *Pfsa*1-4 loci

The *Pfsa*1+ and *Pfsa*3+ mutations have previously been shown to exhibit evidence of association to HbS in the severe cases from Gambia and Kenya, as well as mild cases from Ghana and Mali [20, 22]. We noted that the *Pfsa1* and *3* associations with HbS were extremely strongly supported in this meta-analysis ($BF_{avg} > 1\times10^{29}$; **Figure 2A**, **Figure S4**, and **Supplementary Table 2**).

Two nearby non-synonymous SNPs at the *Pfsa*3 locus, chr11:1,057,437:T>C and chr11:1,058,035:T>A, exhibited the strongest and second strongest evidence for association with HbS genome wide with very large effects (*OR*=24.34 (14.670-40.374) and *OR*=25.11 (15.056-41.893)). Although the latter variant (chr11:1,058,035:T>A) was identified as the most HbS-associated in previous analyses [20, 22], chr11:1,057,437:T>C was notable because of its proximity to a

*Plasmodium* export element (PEXEL)-like motif, suggesting a potential mechanism of effect through an alteration to protein transport [22]. Our analysis adds further weight to the importance of this site, however it is worth noting that structural variation has also been identified at this locus which complicates the fine-mapping of the association signal [22].

The next strongest associations were observed at the *Pfsa*1 locus. Unlike within the discovery analysis, the most-HbS associated variant at *Pfsa*1 was the non-synonymous variant chr2:631,092:T>C, followed by chr2:630,817:G>T (**Supplementary Table 2).** The latter site was not included in the *Pfsa* discovery analysis because of the presence of a second rare alternate allele (chr2:630,817:G>A), which caused it to be filtered from tested variants. In our analysis, the rare G>A allele was removed from the analysis, while the G>T allele was retained (samples carrying the G>A allele were considered to be missing). Both alleles are present within a MSA of a set of whole genome assemblies [18], indicating that this site is truly multiallelic rather than the result of a sequencing or calling error. The site that was originally identified at the *Pfsa*1 locus, chr2:631,190:T>A, was the fifth strongest association with HbS genome wide. The multiallelic site chr2:630,817:G>T is therefore a potential functional candidate to explicate the association of *Pfsa*1 to HbS.

Inspection of forest plots (**Figure S16**) demonstrates that these associations have similar strength in most of the component populations; however, the estimated effect for *Pfsa1+* in Gambia is notably lower than in other sample sets. The allele frequency of *Pfsa*1+ and *Pfsa*3+ alleles was much greater in the CP1 Gambian samples compared to within the GAMCC cohort (**Figure S8A**). This pattern of increasing allele frequency has also been noted within historical samples from the Gambia collected in the 1960s, which may be suggestive of a shift in evolutionary forces within the Gambian population [39]. Therefore, although strength of evidence and effect size at the *Pfsa*3 locus is greater than that of the *Pfsa*1 locus, suggesting a potentially more important relationship between *Pfsa*3 and HbS, it is possible that this has been confounded by a yet unclear evolutionary process within recent samples collected in the Gambia, leading to marginally decreased prominence at the *Pfsa*1 locus.

*Pfsa2+* is only present in east Africa and so this analysis does not add any new information to the previous analysis [1]. In Kenya, the effect of this locus was strong (*OR*=17.77 (8.338-37.887), but strength of evidence in the current meta-analysis is comparatively diminished due to the lack of evidence of association within a small number of *Pfsa*2+ samples present in the Gambian CP1 collection **(Fig. S16).** Overall, this suggests an important role of *Pfsa*2 in East Africa, but further datasets are required to validate this.

The previously implicated *Pfsa4+* variant (chr4:1,124,172 T>A), which lies in *FIKK4.2*, was also associated in our meta-analysis but at lower levels of evidence ($BF_{avg}$=1.41×$10^7$). This variant is only polymorphic in West African populations and support for association comes from the Ghana and Mali datasets (**Figure S16**) [20]. For unknown reasons, the estimated association is notably weaker in both Gambian datasets despite being at high frequency there (the frequency of the sickle-associated allele ($f^{Pfsa4+}$)= 9.7% in GAMCC; $f^{\text{Pfsa4+}}$=16.4% in Gambian CP1 samples).

#### Assessment of independence of association and linkage disequilibrium between *Pfsa* loci on chromosome 2 and 3

Our meta-analysis highlights two clusters of HbS-associations on the same chromosome within the parasite genome. One of these falls on chromosome 2, which carries a ~250Kb region containing the previously identified *Pfsa1, Pfsa2* loci, and a novel putative association in an uncharacterised gene (*Hyp2*) (**Fig. 2B).** The second comprises of multiple novel putative associations within a ~61Kb region on chromosome 3 in the *FIKK3, PTP7* and *CLAG3.1* genes (**Fig. 2C).** These regions are large, and recombination within the *P. falciparum* genome is thought to rapidly decay within ~500bp [40], however we observed that genotypes at these loci are correlated with one another within our datasets (**Fig. S6**). We therefore wanted to assess the possibility that the associations on each chromosome are driven by one causal site that is physically linked to each associated site.

To test whether these represent independent signals of association we took two approaches: (i) conditional analysis and (ii) linkage disequilibrium assessments, the results of which for each region will be detailed below.

**Chromosome 3 loci**

*(i) conditional analysis*

To perform conditional analyses on the chromosome 3 region, we used genotypes at associated loci in the *CLAG3.1* and *FIKK3* genes as covariates and reperformed the analysis. The association at *CLAG3.1* did not appear impacted when conditioned on *FIKK3* genotype, however the signal at *FIKK3* was reduced when conditioned on *CLAG3.1*. The association at *PTP7* was not affected by association at either chromosome 3 locus (**Fig. S17**).

*(ii) linkage disequilibrium estimates*

We calculated Pearson's correlation coefficient (R) and D' and between the *FIKK3* locus (chr3:79,845 G>A) and all biallelic variants in our analysis set between chr3:70,000-150,000 (with an allele frequency > 5%). By plotting R values, we observed that correlation appeared to decay with genomic distance between all sites (**Fig. S7B** shows absolute R value averaged within an 100bp window**)**. We identified that the absolute R and D' values were moderate between *FIKK3* and the other loci (|r| ~ 0.18–0.54, |D′| ~ 0.44–1.0), but were lower across frequency matched sites in the region (average |r| < 0.07, average |D′| ~ 0.205-0.54) (**Supplementary Table 6)**.

Together this suggest that the region is unlikely to form a contiguous haplotype block encompassing all associated sites, however whether these loci represent truly independent associations to HbS, or are in some other way co-associated, remains to be deciphered.

**Chromosome 2 loci**

*(i) conditional analysis*

As described above, we re-performed the analysis across chromosome 2 but using the *Pfsa1* genotype as a covariate. In doing so, signals at both the *Pfsa2* and *Hyp2* loci largely disappeared (**Fig. S18)**.

In addition, we expanded this analysis to all previously identified and novel putative *Pfsa* sites and included *Pfsa3* as an additional covariate. Although the effects at *Pfsa*1 and *Pfsa*3 did not disappear when cross-conditioned on each other, they were reduced (**Fig. S18**). Signals at all other sites were reduced to near no effect when conditioning on both alleles apart from a locus in *CLAG3.1* and *REX2* (**Fig. S18**). Although many of these HbS-*Pfsa* associations therefore do not appear statistically independent of one another, they are connected through prominent between chromosome LD which is challenging to explain without a biological relationship between them which is related to HbS.

*(ii) linkage disequilibrium estimates*

R values to the *Pfsa1* site across the chromosome 2 region clearly decayed between this site and the region containing the *Pfsa2* and *Hyp2* site across all populations (**Fig. S7A).** *Pfsa2* and *Hyp2* are more closely clustered at ~10Kb apart, however they have opposing spatial distributions where *Pfsa2* is absent in West-African populations. Based on this analysis, we observed that the site in *Hyp2* was strongly correlated to a site nearby to *Pfsa2*+ (chr2:814,297 C>T) which is within the same gene as *Pfsa*2+ (*PF3D7_0220300*) (highlighted on **Fig. S7A).** This site is also polymorphic in Kenya, but by a different substitution (C>A). The evidence of association to HbS in our data however was modest (chr2:814,297 C>T; $BF_{avg}$ =345.3, *P*=2.59×10$^{5}$).

In **Supplementary Table 7** we report absolute R and D' between *Hyp2* and each other locus. We observed moderate correlation between *Hyp2* and the additional site in *PF3D7_0220300* across datasets (|r| = 0.56-0.80, |D'|=0.72-0.85). Correlation between *Hyp2* and *Pfsa*1+ and *Pfsa*2+ was less pronounced (|r|~ 0.28-0.49 where *Pfsa*2+ is polymorphic at a moderate allele frequency. We also computed the mean correlation across all sites between *Pfsa*1+ and the *Hyp*2 site (excluding the focal loci) that have a similar frequency to the *Hyp2* site which was low (|r| < 0.065, |D'| < 0.242).

These results suggest that the *Hyp2* site is not physically connected to the *Pfsa1+* mutation but may be part of a small cluster of local LD near to *Pfsa*2, that is correlated with the *Pfsa1+* mutation across East and West African populations.

### Supplementary Figures

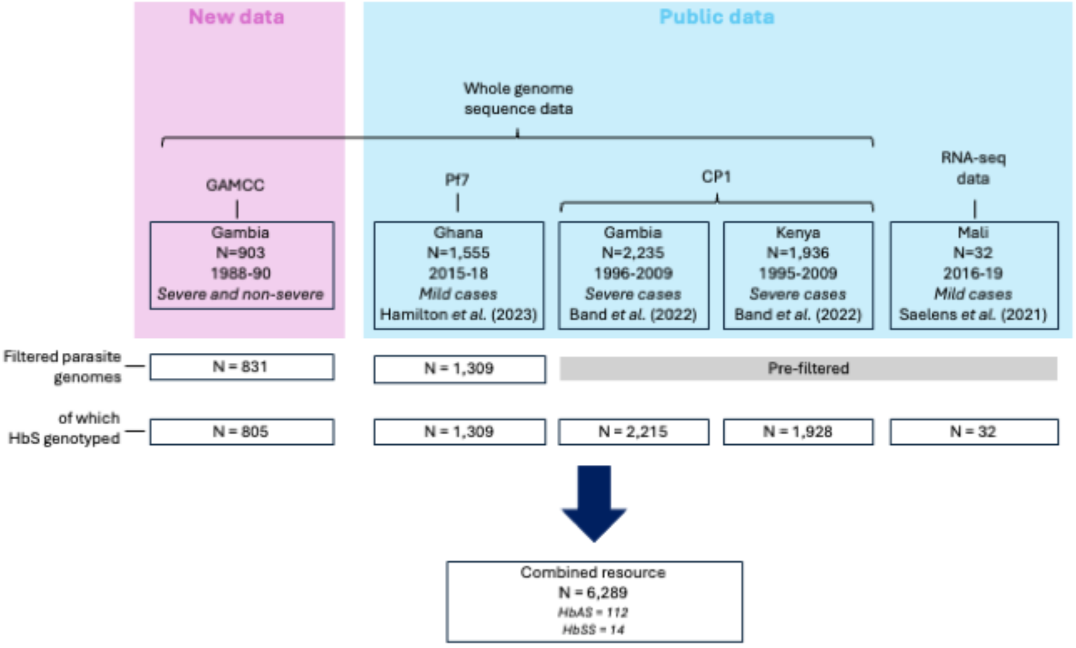

**Figure S1 – Available datasets for analysis of HbS-associations within the *P. falciparum* genome**

Flowchart showing the generation of a data resource to assess for associations between HbS and *P. falciparum* variation using 5 datasets from 4 populations in Africa.

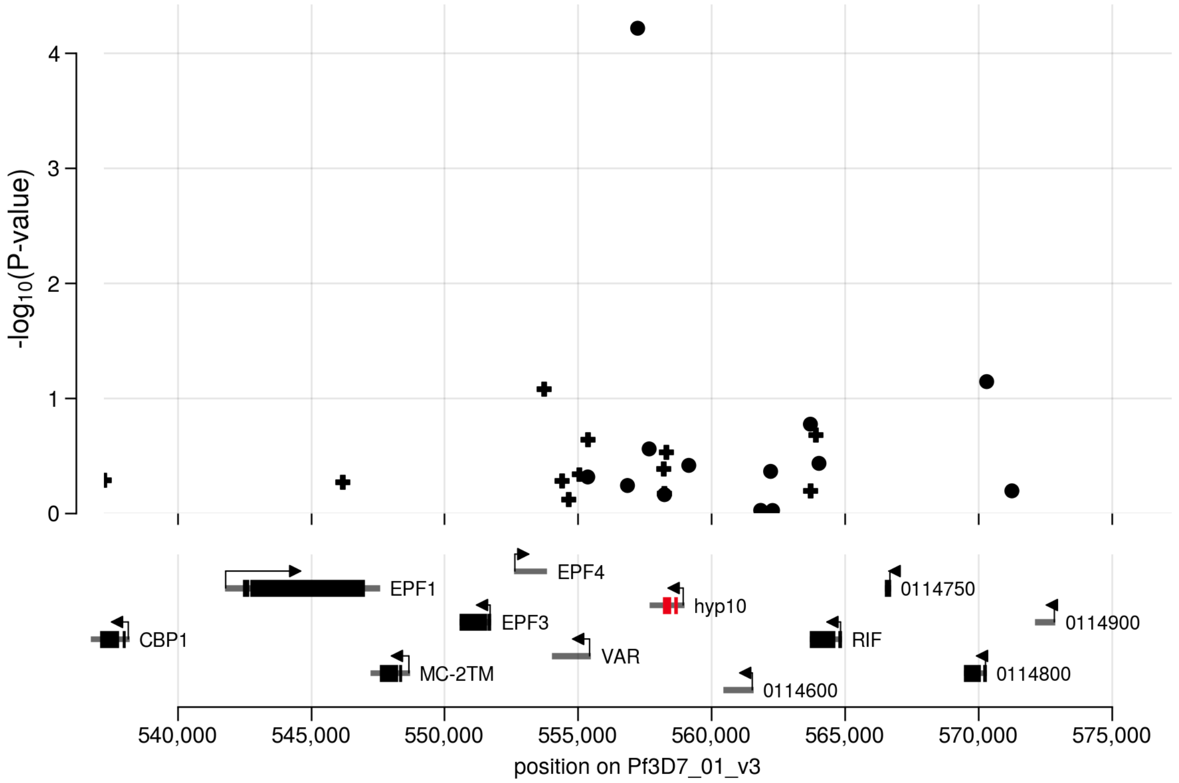

**Figure S2 – Detail of chr1 association with severe vs. non-severe disease in the GAMCC dataset**

A detailed view of the putative severity locus at chr1:557,240 T>A identified in a test for association between parasite variation and severe or non-severe disease in the GAMCC dataset. A 40Kb window flanking the associated site is shown. Biallelic core variants with <10% missing threshold and a 2.5% minor allele frequency within the GAMCC are shown. The position of genes at the locus is shown at the bottom of the plot and the gene containing the putative severity site (*PF3D7_0114500*, *hyp10)* is highlighted in red.

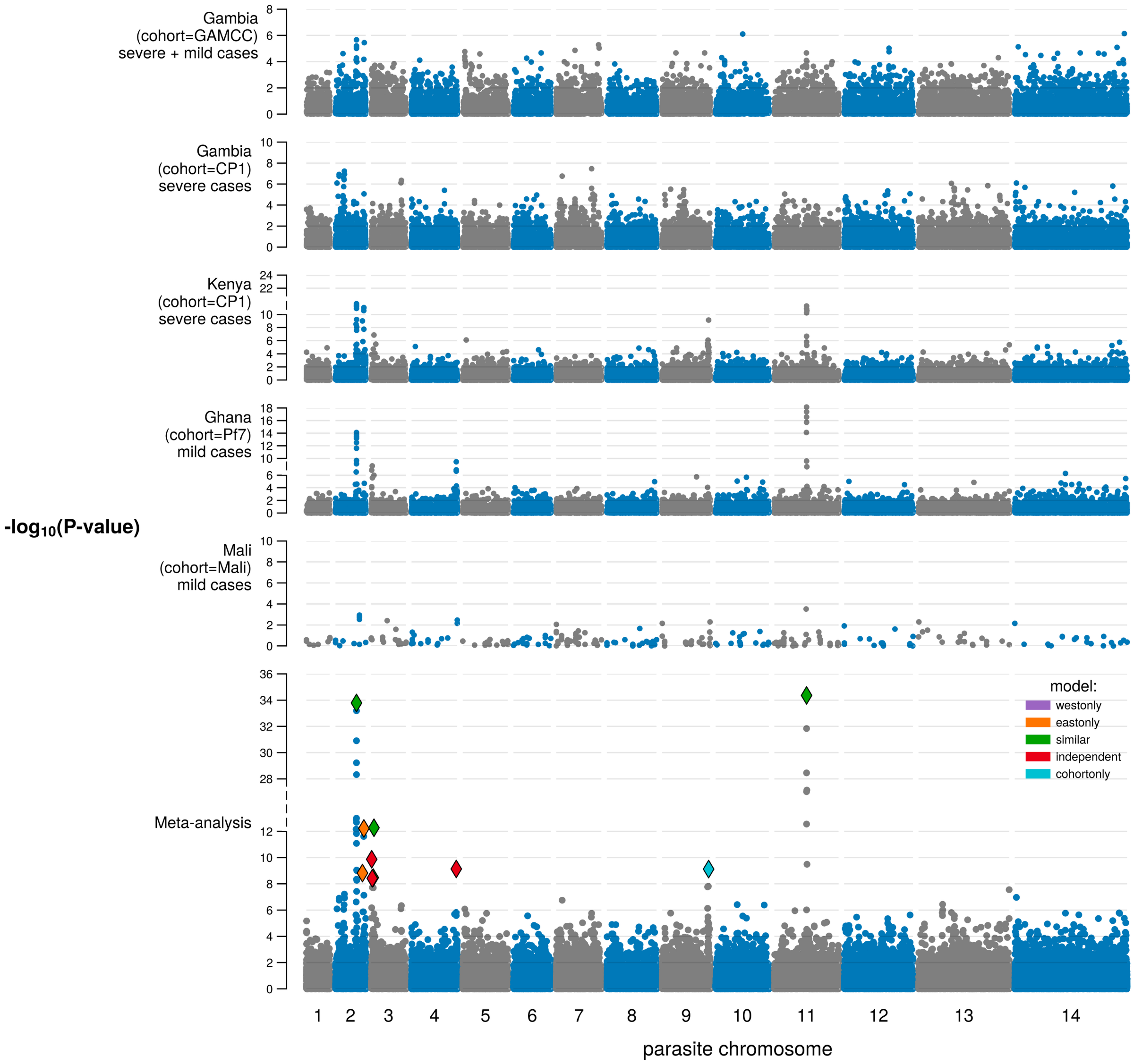

**Figure S3 – Frequentist fixed effect meta-analysis of association with HbS**

Frequentist fixed effect meta-analysis of HbS-associations across individual datasets. Each row shows results of a frequentist association test between HbS genotype as the predictor and P. falciparum variation as the outcome. Only biallelic, core variants with <25% of calls missing and an allele frequency of at least 1% across the combined dataset are shown. Model of highest evidence is indicated by a coloured diamond on the meta-analysis plot to allow comparison with Figure 2 in the main text.

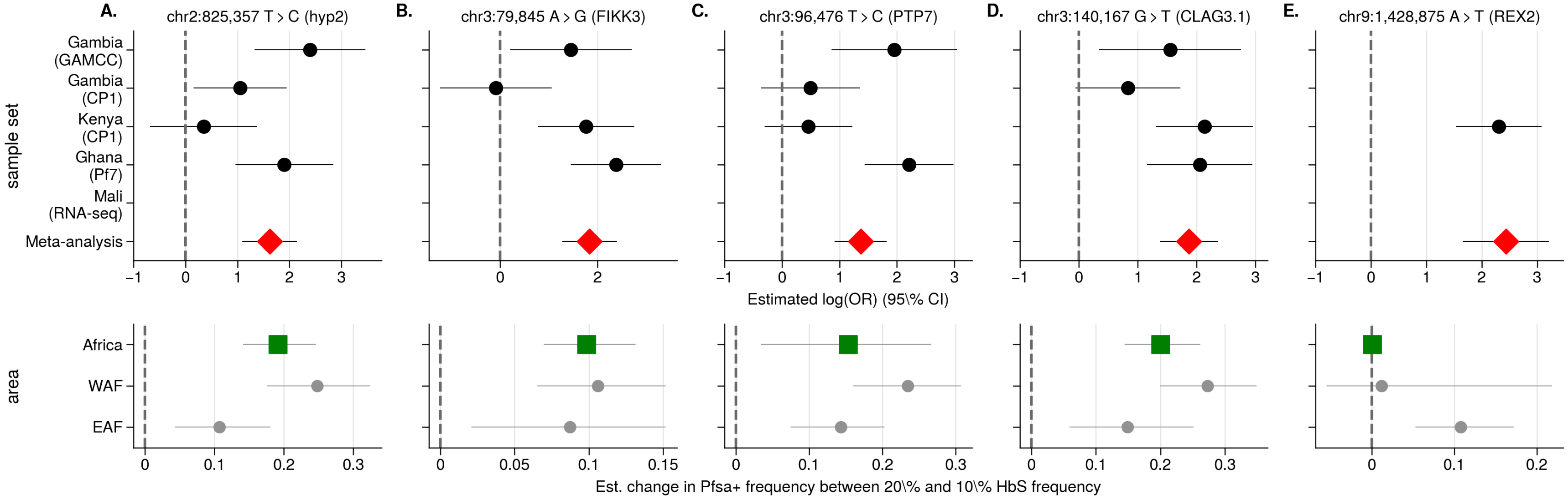

**Figure S4- Combined association and geographic evidence at novel putative *Pfsa* loci**

Evidence for association with HbS across datasets. The estimated effect of HbS on parasite genotype at each site (x-axis) as a logOR with 95% confidence intervals is shown. These estimates were computed using a logF(2,2) prior. The meta-analysis effect size is indicated as the bottom row. This was computed by running associations individually within each dataset using an uninformative prior (logF(0.1,0.1)) and meta-analysing across datasets comparing different models of effect as described in the main text. Variant calls at these sites were not available for the Mali dataset and so estimates are missing from the plots. Below shows results from a recent geospatial analysis of HbS and *P. falciparum* allele frequency correlation across Africa. The x-axis shows the change in frequency of the parasite variant across an increase in HbS frequency from 10% to 20%.

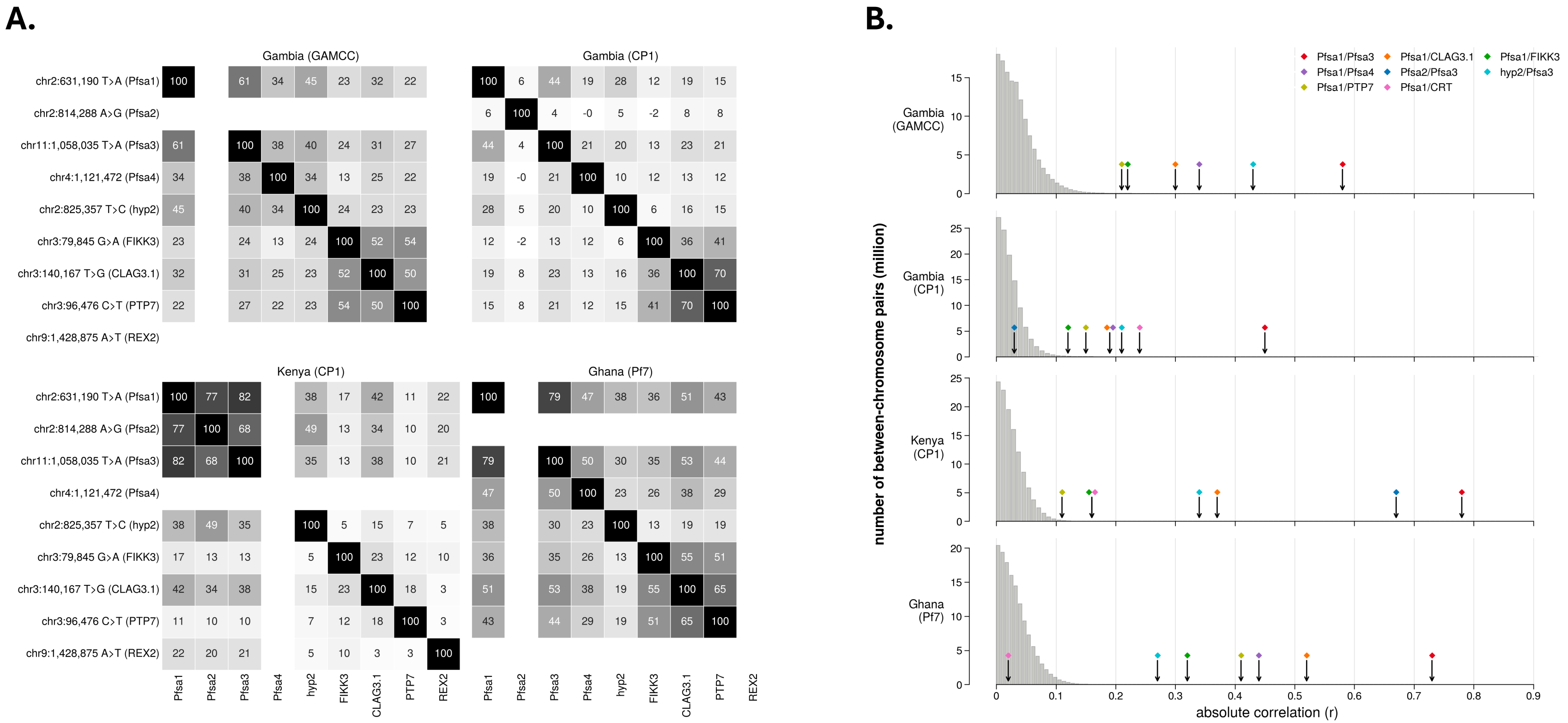

**Figure S5 – Positive linkage disequilibrium between known *Pfsa* loci and putative novel *Pfsa* loci in four datasets**

**A**.Pearson's corelation coefficient R (%) between *Pfsa*1-4 and putative *Pfsa* loci within the four available whole genome sequence datasets. Higher values of correlation are also indicated by a deeper grey colour. Where a site was completely missing (allele frequency of <1%) cells are left blank. Sites have been orientated to reflect *Pfsa*+ alleles as 1and *Pfsa*- alleles as 0. **B**. Each row shows the distribution of between chromosome linkage disequilibrium between all pairs of biallelic variants (with a minimum allele frequency of 2.5% and a minimum sample size of 500) within each dataset indicated on the y-axis. Coloured diamond markers with arrows show the linkage disequilibrium value (absolute Pearson's R) between a previously identified Pfsa locus on a different chromosome (chr2:631,190:T>A (Pfsa1) or chr11:1,057,437 T>C (Pfsa3)) and another locus of interest, as indicated in the legend.

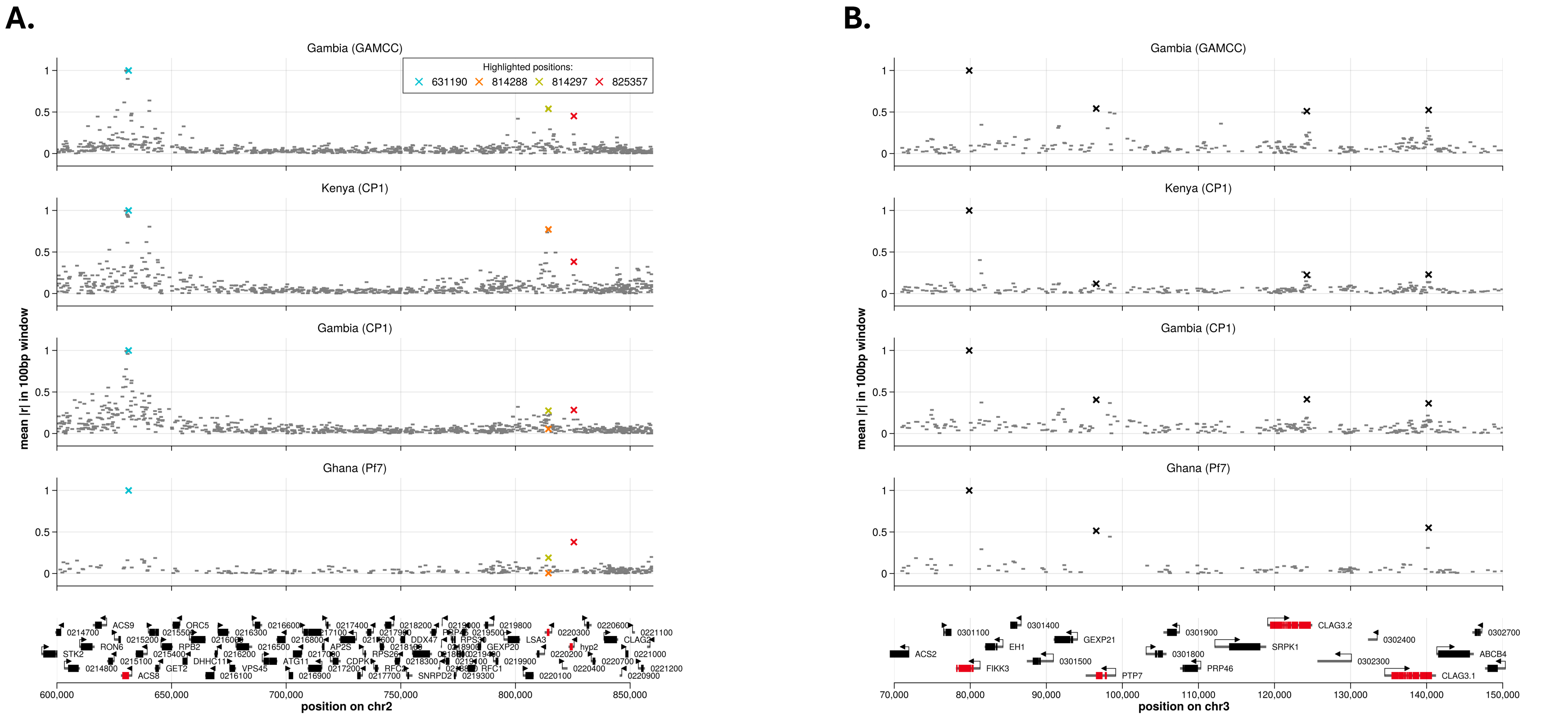

**Figure S6 - Local linkage disequilibrium (R) between nearby known and novel putative *Pfsa* variants**

Correlation (Pearson's R) to leftmost known/putative *Pfsa* variant (**A**. chr3:631,190 T>A (*Pfsa*1) and **B**. chr3:79,845 G>A) with all other variant sites (allele frequency > 5%, sample size > 500) between: **A**. chr2:600,000-860,000 and **B**. chr3:70,000-150,000, averaged within 100bp windows in each dataset. Sites of association to HbS within the region are denoted with 'X' markers (in **A**. chr2:631,190 T>A (*Pfsa*1), chr2:814,288 C>T (*Pfsa*2) and *Pfsa*+ mutation chr2:825,357 T>C, as well as a site which we noted was also correlated with *Pfsa*1 in the GAMCC dataset (chr2:814,297 C>T, see **Supplementary Info**), and in **B**. chr3:79,845 G>A (in the FIKK3 gene), chr3:96,476 C>T (PTP7), chr3:140,167 T>G (CLAG3.1) and its paralogous site in CLAG3.2 (chr3:124,242 T>G)). For clarity, sites in **A** have been coloured as indicated in the legend to more easily allow their identification. The positions of genes across the region are shown at the bottom of the plot, and those containing sites of association to HbS have been highlighted in red.

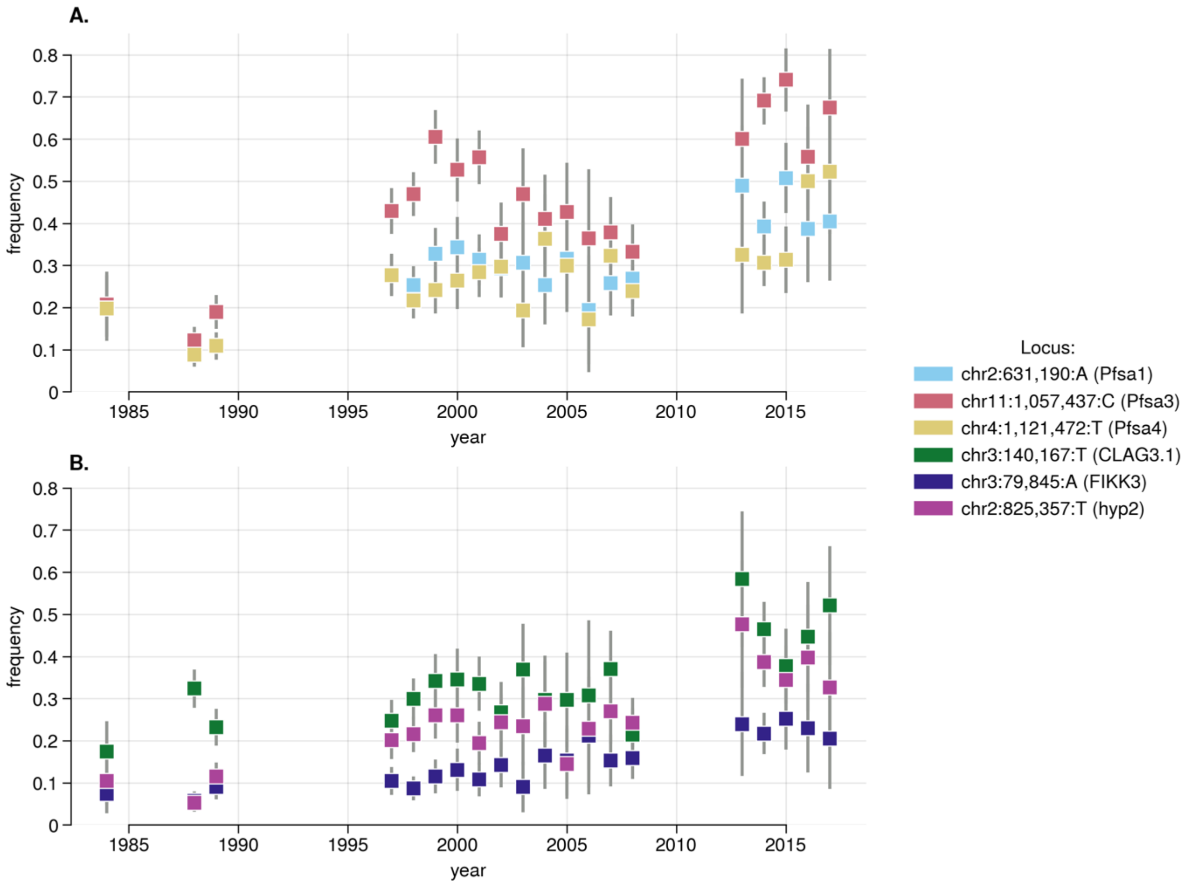

**Figure S7 – Temporal change in allele frequency at known and novel putative *Pfsa* loci in the Gambia**

Change in identified *Pfsa* (top) and novel putative *Pfsa* (bottom) loci allele frequencies (y-axis) in data collected in The Gambia by collection year (x-axis). The plotted locus is indicated by marker colour. 95% confidence interval error bars are shown. Only years with at least 25 samples are shown.

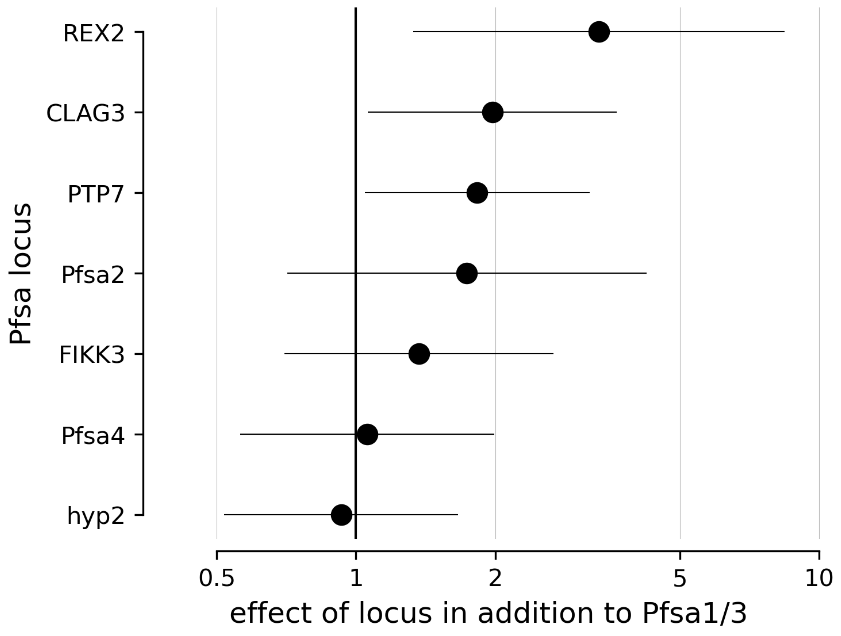

**Figure S8 – Estimated multiplicative effect of putative *Pfsa* loci on the *Pfsa*1/3 background.**

For each previously identified (*Pfsa2* and *Pfsa4*) and putative *Pfsa* locus (y-axis), the additional impact of the *Pfsa*+ allele on the relative risk of infection due to HbS against a background carrying the *Pfsa1*+ and *Pfsa3*+ genotype is shown. Effects were estimated using a multinomial logistic framework whereby the log-RRR parameter for each multi-locus genotype was decomposed into contributions from Pfsa1 and *Pfsa*3, and from the specified locus, as described in Methods. Points denote β effect estimates, and segments show 95% confidence intervals.

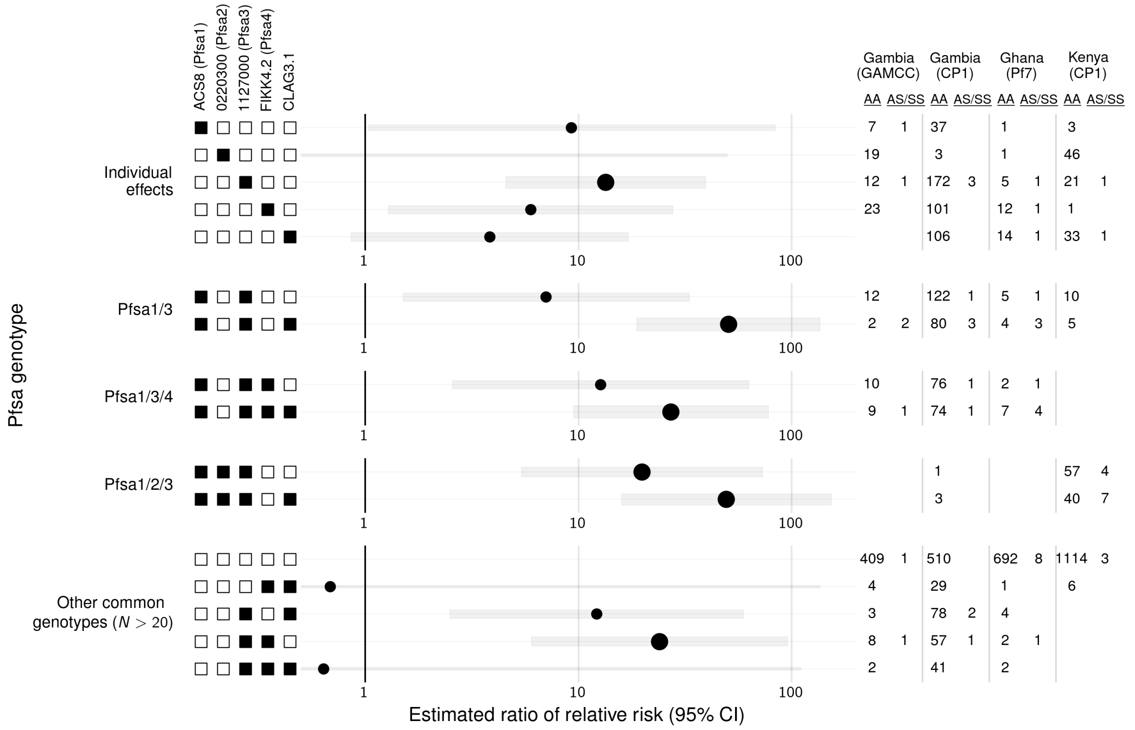

**Figure S9 – Estimated ratio of relative risk (RRR) across different *Pfsa* genotype combinations across all datasets**

Estimated ratio of relative risk of malaria based on HbS for different *Pfsa* genotype combinations (indicated by filled black squares on the left) compared to infections of *Pfsa*- parasites. This analysis was conducted across all available whole genome sequence data with paired HbS genotyping (GAMCC, CP1 Gambian and Kenyan samples and Ghanaian samples) using genotypes at the *Pfsa1* (chr2:621,190 A>T in the ACS8 gene), *Pfsa2* (chr2:814,288 C>T (0220300)), *Pfsa3* (chr11:1,058,035 T>A (1127000)), *Pfsa4* (chr4:1,121,472 A>T (FIKK4.2)) and a putative novel *Pfsa*+ mutation in the CLAG3.1 gene (chr3:140,167 T>G). *RRR*s were calculated using a multinomial logistic regression using *Pfsa*- infections as a baseline. 95% confidence intervals are indicated by the light grey bar. To avoid over-fitting of points with small sample sizes, Bayesian regularisation was performed using a weakly informative Gaussian prior with mean 0 and standard deviation of 10 for each parameter, and between-parameter correlation set to 0. Estimates calculated with four or more HbS individuals, and thus have higher confidence, are indicated by larger points. HbAS and HbSS genotypes were grouped together (i.e. a model of dominance was assumed). To the right, sample sizes stratified by human genotype are shown per dataset.

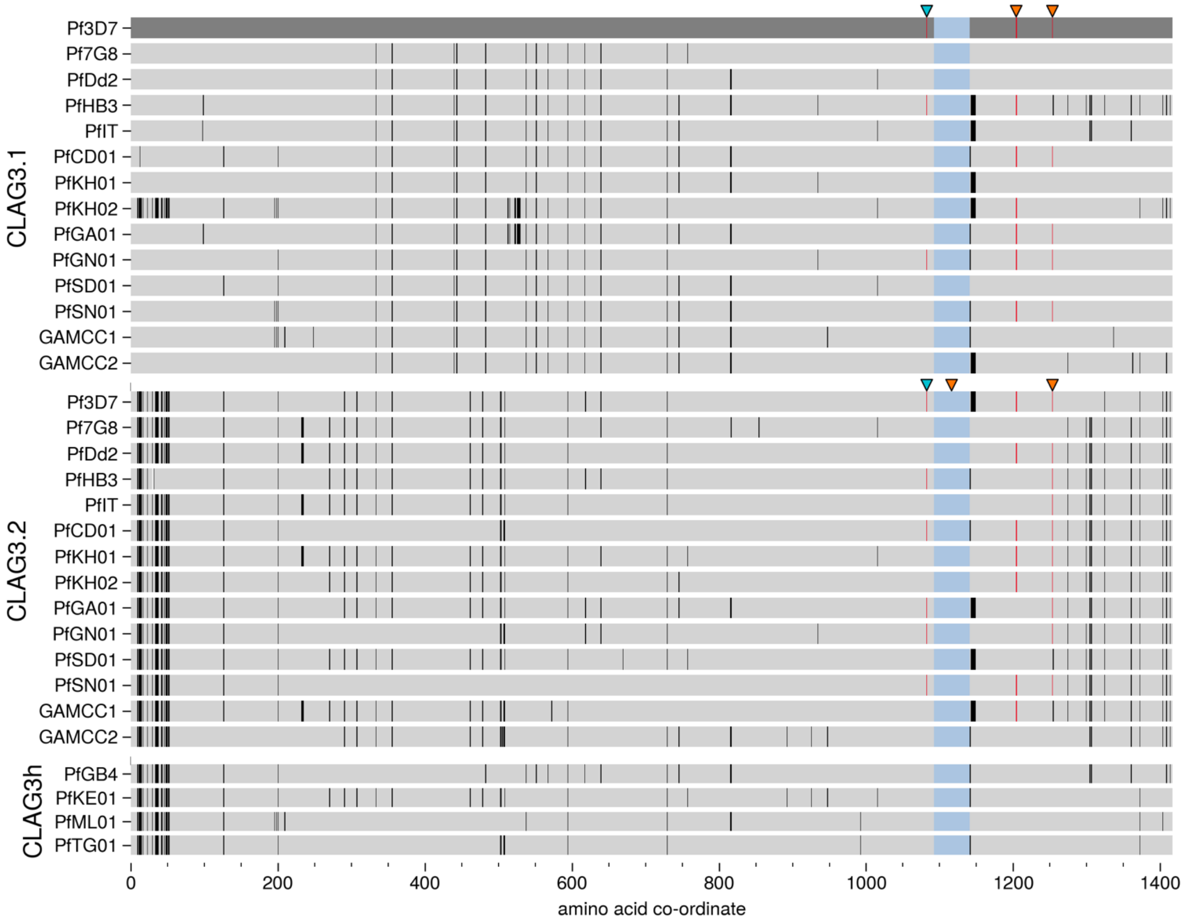

**Figure S10 – Multiple sequence alignment of *CLAG3* gene copies**

A multiple sequence alignment of *CLAG3.1* (top), *CLAG3.2* (middle) and CLAG3h (bottom) amino-acid sequences extracted from a set of 15 long read assemblies [18] and newly generated data for two GAMCC samples. Amino acid differences (in comparison to the CLAG3.2 sequence in the Pf3D7 reference assembly, shown in dark grey) are indicated in black. A known hypervariable region (HVR) has been compressed and highlighted blue. Non-synonymous amino acid substitutions with evidence of association to HbS ($\log_{10} BF_{avg} > 4$ on Figure 2) in each gene are indicated by orange arrows; in CLAG3.1 chr3:140,020 T>C (V1205A) and chr3:140,167 T>G (I1254R), and in *CLAG3.2* chr3:124,095 T>C (V1204A) and chr3:124,242 T>G (I1253R). A blue arrow indicates a site identified as associated with HbS following paralogy-aware variant calling across this locus (Figure 4 in the main text); chr3:123,737 C>A (L1085I) in CLAG3.2 and chr3:139,653 C>A (L1083I) in *CLAG3.1*. Where a *Pfsa*+ allele was indicated to be the reference allele in Pf3D7 *CLAG3.2*, the highlighting of differences has been switched such that the *Pfsa*+ allele is shown (as indicated using the colour red).

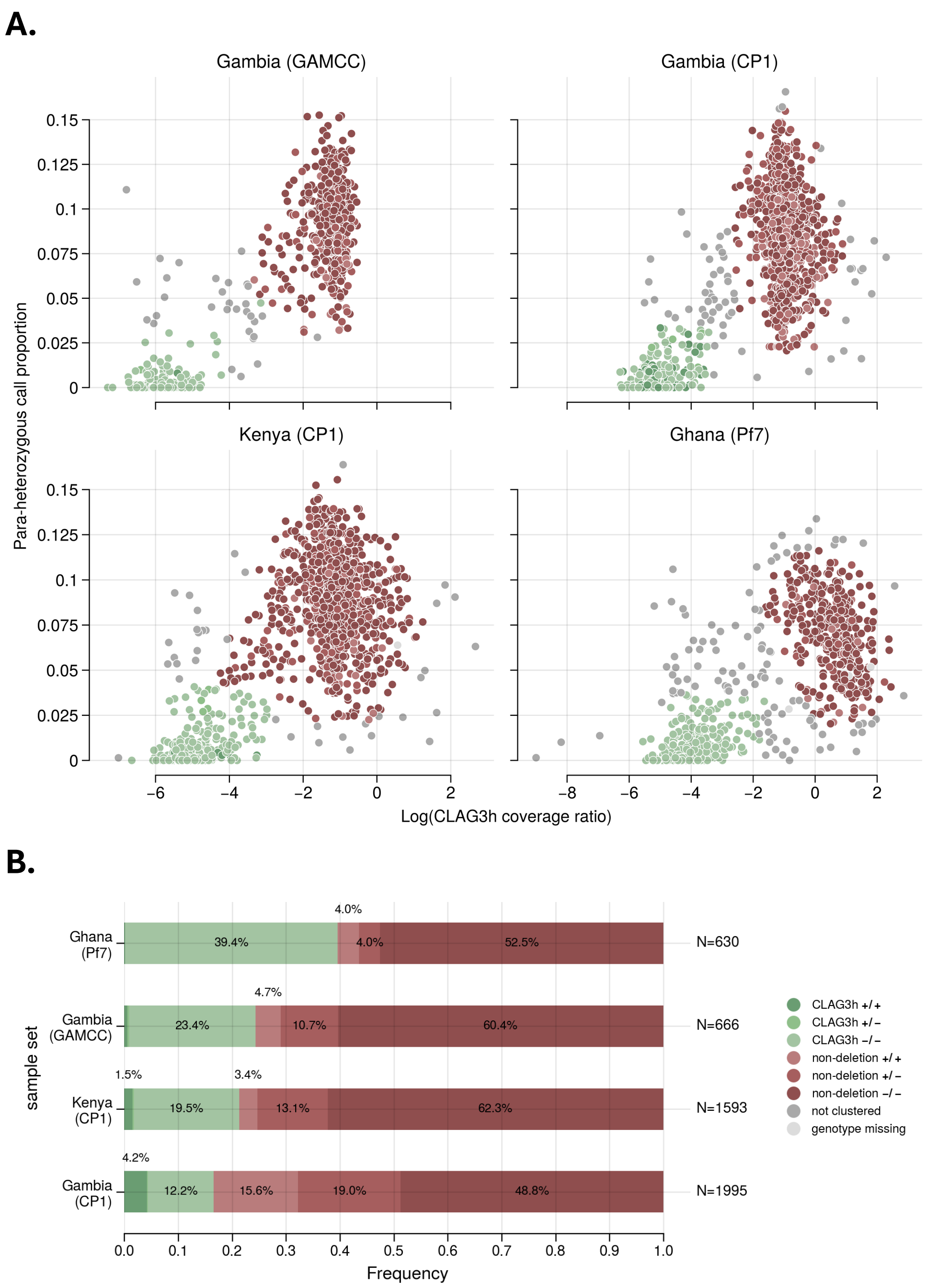

**Figure S11 – Results of CLAG3h calling across datasets**

**A.** Points show CLAG3h call for samples within each dataset. Density clustering was performed to call presence of the CLAG3h structural variant at the *CLAG3* locus in each dataset. Clustering was performed using the logarithm of the coverage ratio (x-axis), calculated by comparing coverage between the CLAG3.1 and CLAG3.2 to flanking regions within alignments to the 3D7 reference assembly, and using proportion of para-heterozygous calls in the renewed variant calling across the *CLAG3* locus (y-axis). **B.** Frequency of CLAG3h and samples carrying two *CLAG3* gene copies within each dataset. Sample size and frequency value are also shown. Only categories with more than 1% frequency are labelled. As indicated by the legend, CLAG3h is marked by green and samples carrying two CLAG3 copies in red, and each has been further stratified to indicate frequency of genotypes of the putative *Pfsa* chr3:140,167:T allele.

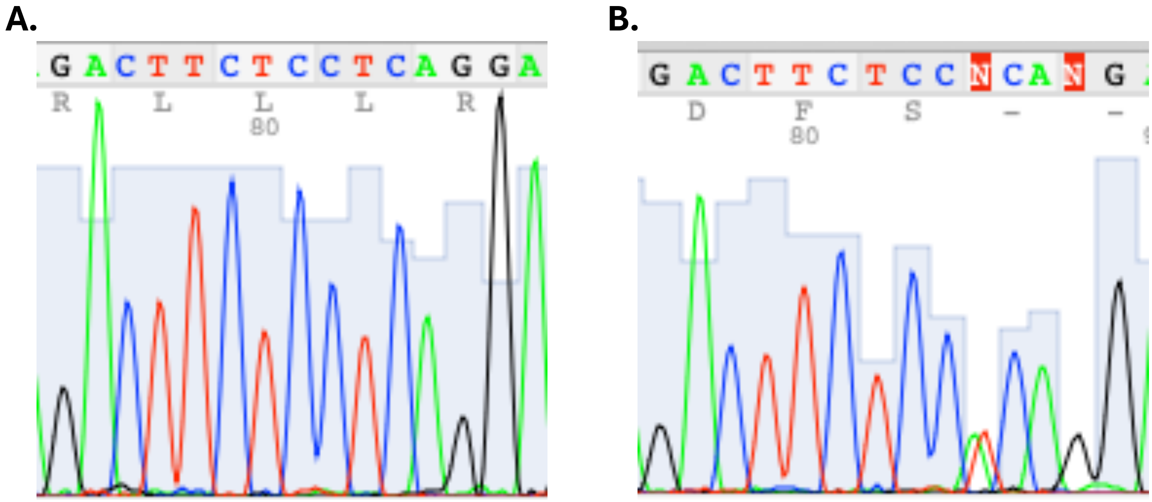

**Figure S12 - Example Sanger-sequencing traces used to genotype the HbS locus.**

Sanger sequencing trace image for A. an HbAA sample and B. an HbAS sample.

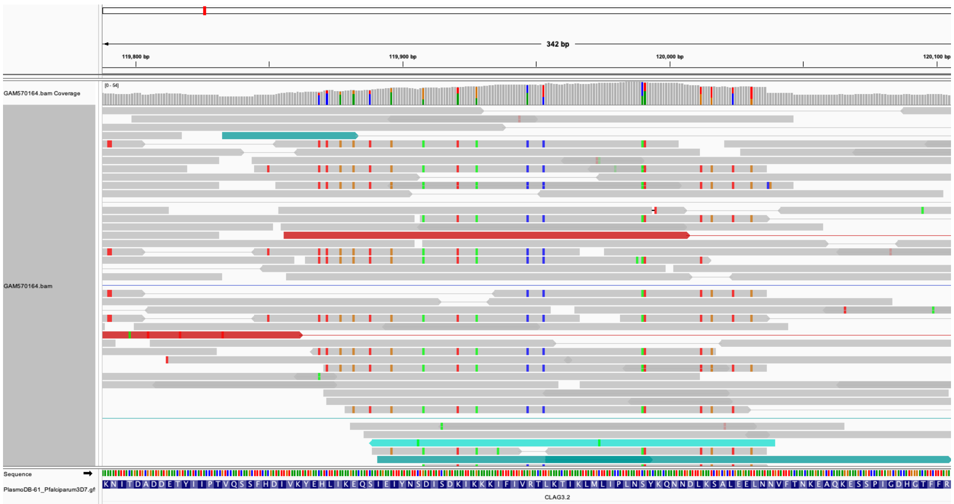

**Figure S13 – Example reads aligning to the CLAG3 region**

An example of reads aligning to the CLAG3 region which show entirely different genotypes to others aligning there. Viewed using IGV.

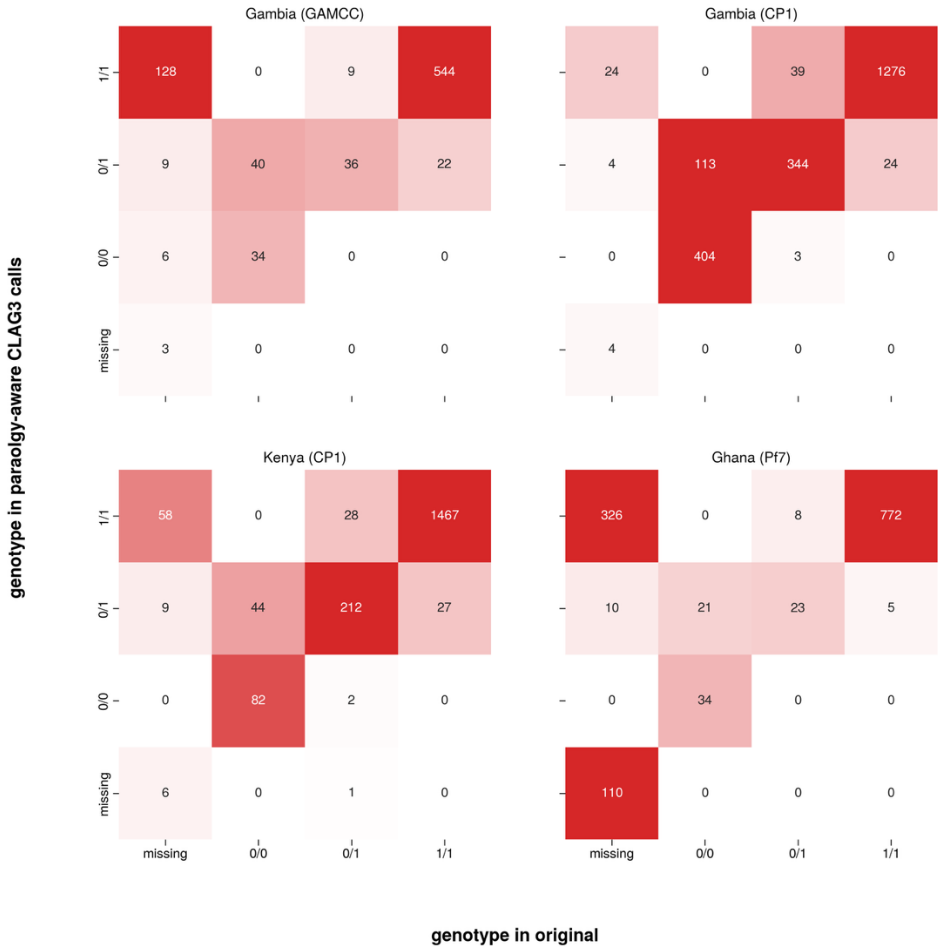

**Figure S14 – Comparison of paralogy-aware and unaware genotype calls at CLAG3**

A matrix showing the change in counts of genotype calls between the original variant calls (columns) and paralogy-aware variant calls (rows) at a putative sickle-associated site within each dataset. In the original vcf thjs corresponds to chr3:140,167 T>G and in the renewed vcf this site is represented as a diploid call spanning the para-polymorphic sites chr3:140,167 and chr3:124,242 T>G.

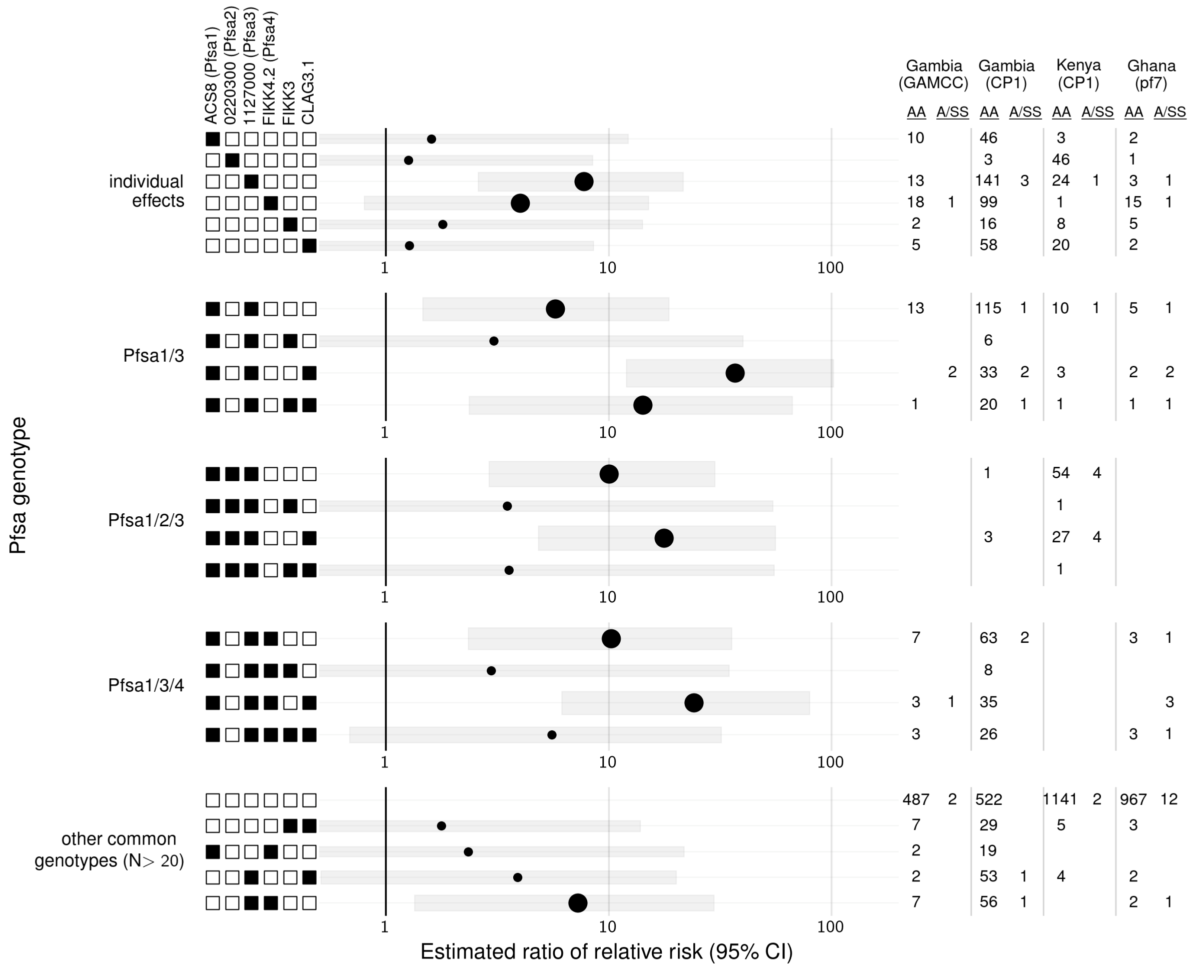

**Figure S15 - Estimated ratio of relative risk of malaria based on HbS for different *Pfsa* genotype combinations**

Figure shows the estimated ratio of relative risk of infection due to HbS (RRR, x axis) for different *Pfsa* genotype combinations (rows, as indicated by filled black squares on the left) compared to infections of *Pfsa*- parasites made using paralogy-aware *CLAG3* calls. This analysis was conducted across all available whole genome sequence data with paired HbS genotyping (GAMCC, CP1 Gambian and Kenyan samples and Ghanaian samples) using genotypes at the *Pfsa*1 (chr2:621,190 A>T in the ACS8 gene), *Pfsa*2 (chr2:814,288 C>T (0220300)), *Pfsa*3 (chr11:1,058,035 T>A (1127000)), Pfsa4 (chr4:1,121,472 A>T (FIKK4.2)) and putative novel *Pfsa*+ mutations in the *FIKK3* (chr3:79,845 G>A) and *CLAG3.1* (chr3:140,167 T>G) loci. RRRs were calculated using a multinomial logistic regression using Pfsa- infections as a baseline. 95% confidence intervals are indicated by the light grey bar. To avoid over-fitting of points with small sample sizes, Bayesian regularisation was performed using a weakly informative Gaussian prior with mean 0 and standard deviation of 10 for each parameter, and between-parameter correlation set to 0. Estimates calculated with two or more HbS individuals, and thus have higher confidence, are indicated by larger points. HbAS and HbSS genotypes were grouped together (i.e. a model of dominance was assumed). To the right, sample sizes stratified by human genotype are shown per dataset.

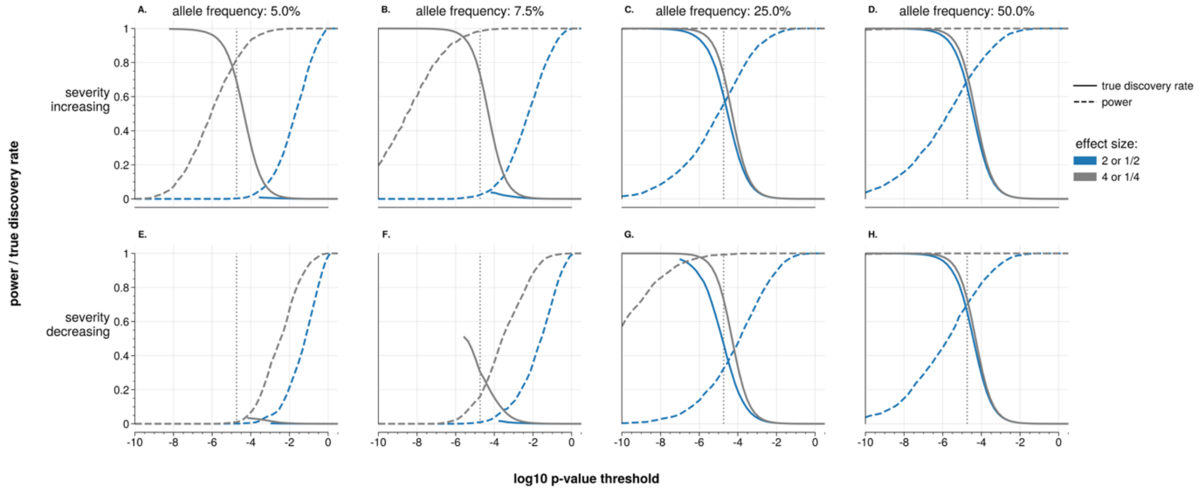

#### Figure S16 - Power and true discovery rate for parasite genetic effects on severity

Power and true discovery rate analysis for an analysis of *P. falciparum* parasite genetic effects on severity using the GAMCC dataset. Model computed power (dashed lines) and true discovery rates (solid lines) for different P-value thresholds (log10) on the x axis. These values were calculated based on 2000 simulations using a prior assumption that 1 in the approximately 20,000 regions in the *P. falciparum* genome is associated with severe malaria. Five baseline allele frequencies (that is, the frequency in mild malaria cases) were assessed: 5%, 10%, 25% and 50% (from left to right). Two effect size magnitudes were used. Severity increasing (**A-D**) indicate the variant increases the chance of severe malaria (i.e. OR = 2 (blue) and OR = 4 (grey)). Severity reducing variants (**E-H**) indicate the variant reduces the risk of severe malaria (i.e. OR = $\frac{1}{2}$ (blue) and OR = $\frac{1}{4}$ (grey)). The grey dashed line represents the Bonferroni correction level (0.05/number of tested variants = $1.8\times10^{-5}$).

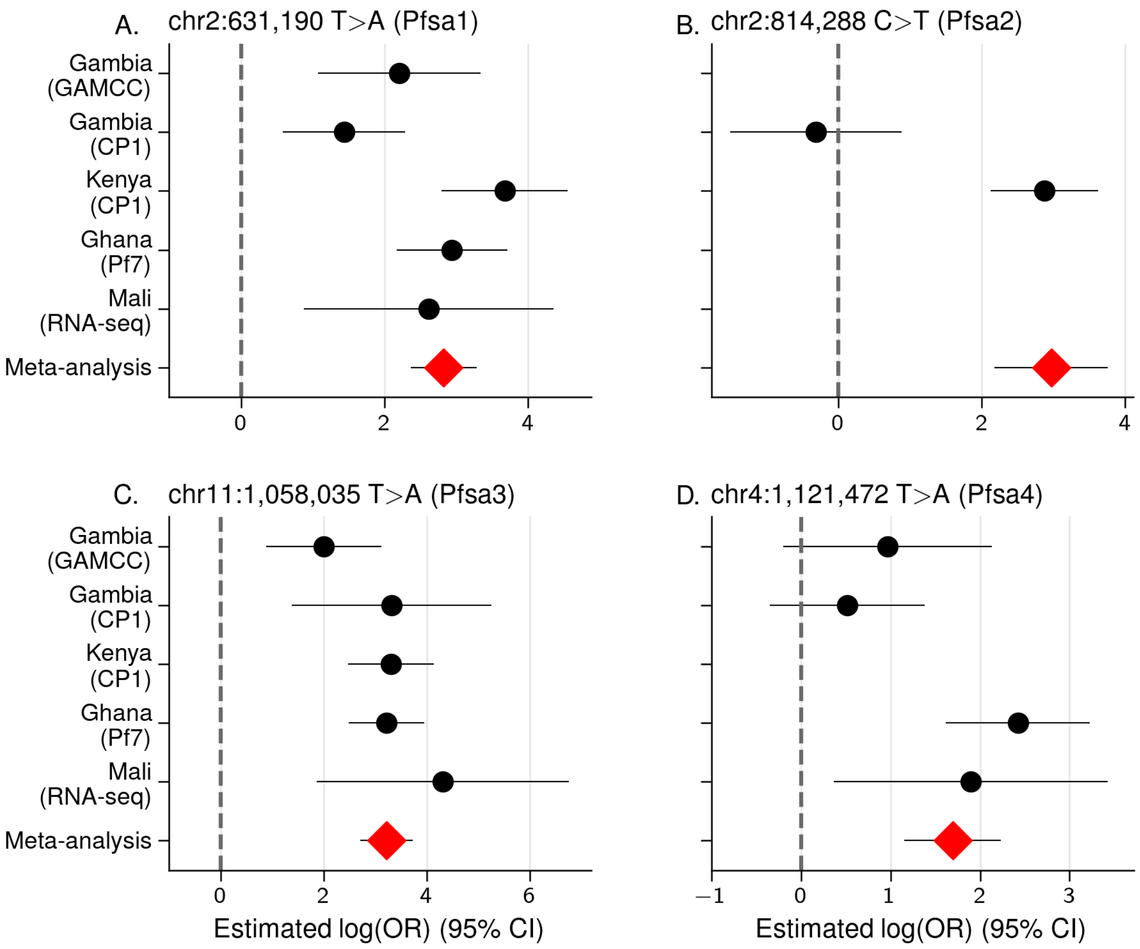

**Figure S17 - Replication of HbS association evidence across the five datasets**

Replication of evidence across the five datasets (indicated on the y-axis), indicating the estimated effect size as a log(odds ratio (OR)) and showing 95% confidence interval (CI)s as lines. These estimates were computed using a logF(2,2) prior. The meta-analysis effect size is indicated as the bottom row. This was computed by running associations individually within each dataset using an uninformative prior (logF(0.1,0.1)) and meta-analysing across datasets comparing different models of effect as described in the main text. If a dataset did not contain the variant due to missingness or a low allele frequency, the effect size is not shown.

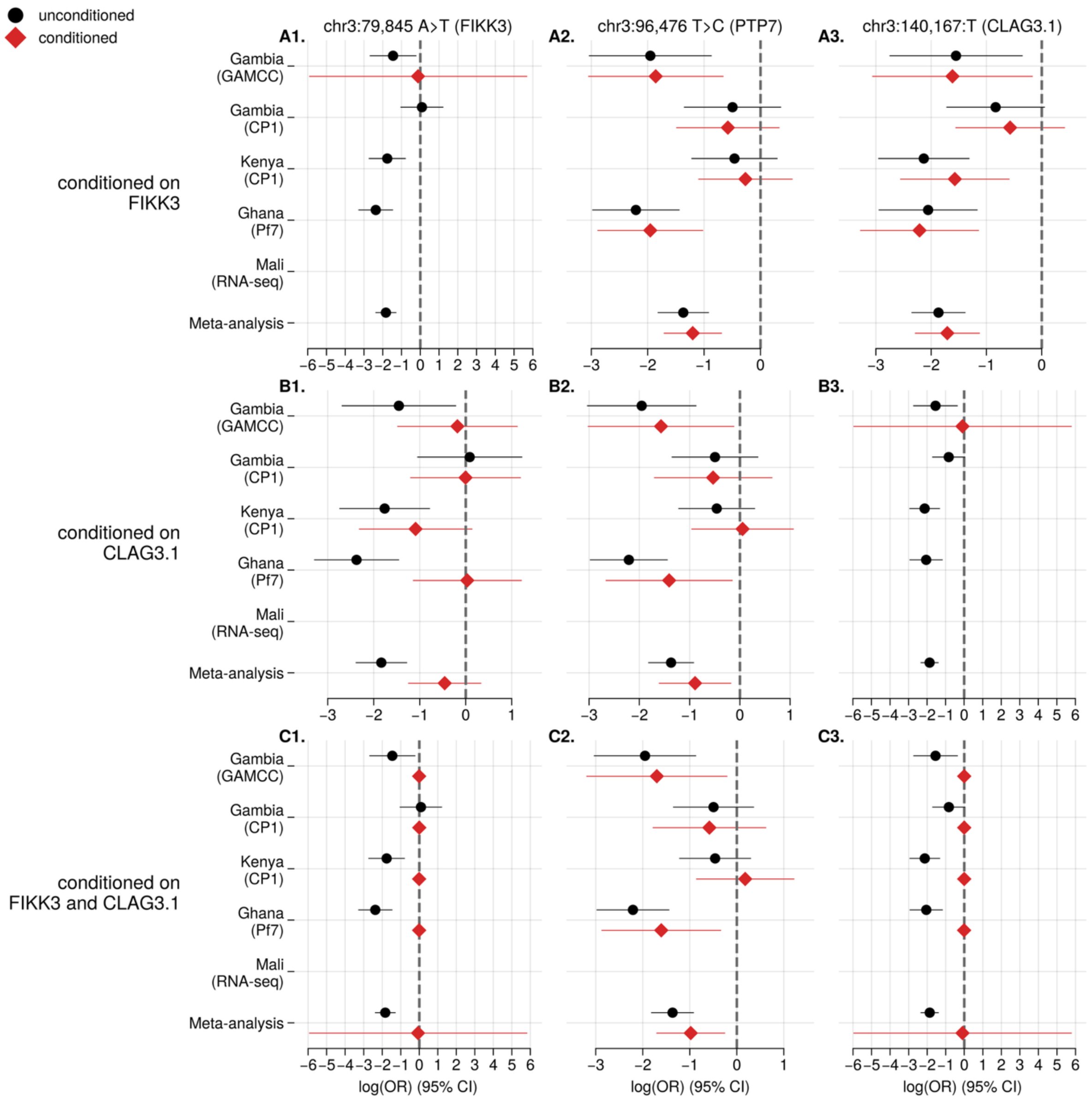

**Figure S18 - Conditional analysis of chromosome 3 *Pfsa* association signals**

The estimated effect of HbS on parasite genotype at a site (1-3) in the analysis presented in the main text (black) and conditional on genotypes at the **A.** chr3:140,167 (*CLAG3.1*), **B.** chr3:79,845 (*FIKK3*) or **C.** both loci included as covariates. Estimates are shown as a logOR with 95% confidence intervals shown.

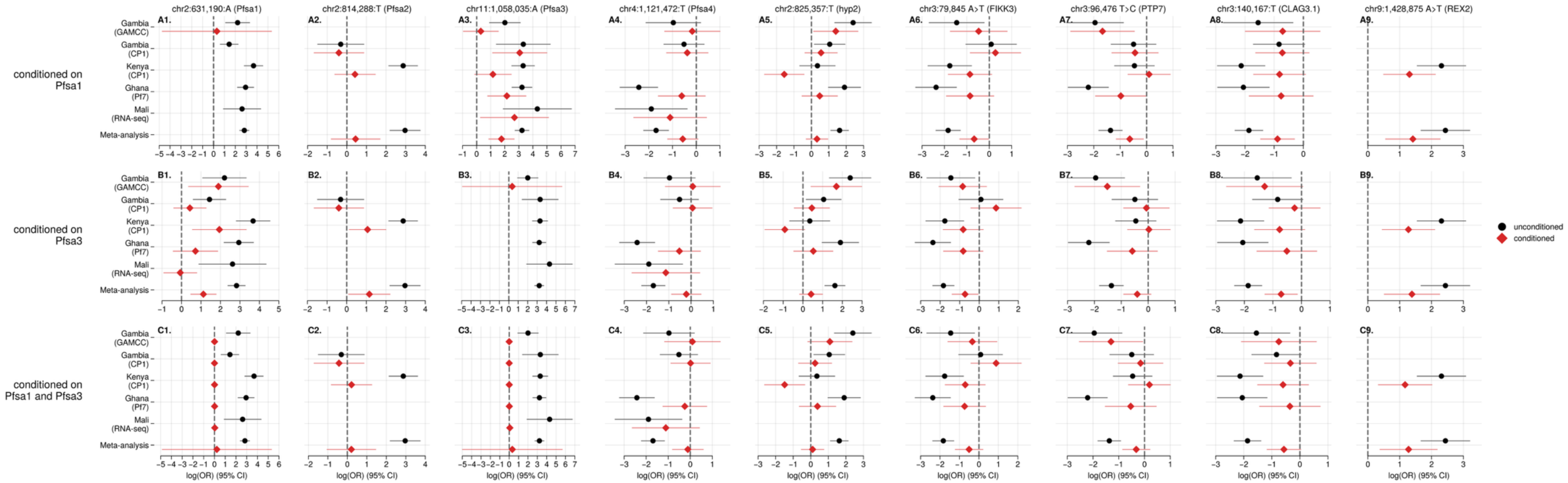

**Figure S19 - Conditional analysis of chromosome *Pfsa* association signals**

The estimated effect of HbS on parasite genotype at each site (1-9) in the analysis presented in the main text (black) and conditional on genotypes at the **A.** chr:631,190 T>A (*Pfsa*1), **B.** chr11:1,058,035 T>A (*Pfsa*3) or **C.** both loci included as covariates. Estimates are shown as a logOR with 95% confidence intervals shown.

### Supplementary Tables

##### Supplementary Tables 1-3

Enclosed within supplementary_tables.xlsx

##### Supplementary Table 4

The PCR program used to generate amplicons within the HBB gene.

| *Step* | *Temperature* | *Time* |
|---|---|---|
| *1* | 96°C | 1 min |
| *2* | 94°C | 45 secs |
| *3* | 56°C | 45 secs |
| *4* | 72°C | 30 secs |
| *5* | Go to Step 2 | x5 |
| *6* | 94°C | 45 secs |
| *7* | 65°C | 45 secs |
| *8* | 72°C | 30 secs |
| *9* | Go to Step 6 | X35 |
| *10* | 72°C | 10 mins |
| *11* | 15°C | 15 mins |
| *12* | END | |

##### Supplementary Table 5

Counts of methods used to call the HbS genotype in the GAMCC dataset. Abbreviations: PCR – amplification of a portion of the HBB gene followed by Sanger sequencing; lcwgs – low coverage whole genome sequencing followed by imputation, chip – array typing on the ThermoFisher Axiom Precision Medicine Array which directly types HbS.

| **HbS genotyping method** | **count** |
|---|---|
| *PCR* | 680 |
| *PCR and lcwgs* | 4 |
| *PCR and chip* | 2 |
| *PCR ,chip and lcwgs* | 15 |
| *chip* | 12 |
| *lcwgs* | 8 |
| *chip and lcwgs* | 93 |
| *not called* | 89 |
| *total* | 903 |

#### Supplementary Table 6

Linkage disequilibrium between the putative *Pfsa+* locus in the *FIKK3* gene (chr3:79,845 G>A) and other putative *Pfsa* loci on chromosome 3, shown separately for each dataset. For each pair, the minor allele frequency (MAF), the absolute Pearson's correlation coefficient (|*r*|) between genotypes, and the absolute D′ value is reported. The final row for each dataset shows the average statistic calculated using all sites in the specified range chr3:79,845-140,167) (excluding the four focal *Pfsa* loci) whose MAF is within ±5% (and with a minimum MAF of 2.5%) of the MAF at the *FIKK3* site in that dataset. "Number of sites" refers to the number of SNPs contributing to each computed value.

| *Position (range)* | *Minor allele frequency (range)* | *Absolute **Pearson's R** to chr3:79,845 G>A (FIKK3) (across range)* | *Absolute **D'** to chr3:79,845 G>A (FIKK3) (across range)* | *Number of sites* |
|---|---|---|---|---|
| | | | | *Gambia (GAMCC)* |
| *chr3:79,845 G>A (FIKK3)* | 11.7% | 1.000 | 1.000 | 1 |
| *chr3:96,476 C>T (PTP7)* | 29.5% | 0.542 | 1.000 | 1 |
| *chr3:124,242 T>G (CLAG3.2)* | 42.7% | 0.511 | 0.751 | 1 |
| *chr3:140,167 T>G (CLAG3.1)* | 31.2% | 0.523 | 0.714 | 1 |
| *chr3:79,845-140,167 (excluding the above)* | 6.7-16.7% | 0.069 | 0.321 | 284 |
| | | | | *Gambia (CP1)* |
| *chr3:79,845 G>A (FIKK3)* | 16.5% | 1.000 | 1.000 | 1 |
| *chr3:96,476 C>T (PTP7)* | 40.7% | 0.407 | 0.762 | 1 |
| *chr3:124,242 T>G (CLAG3.2)* | 40.1% | 0.412 | 0.677 | 1 |
| *chr3:140,167 T>G (CLAG3.1)* | 41.8% | 0.364 | 0.655 | 1 |
| *chr3:79,845-140,167 (excluding the above)* | 11.5-21.5% | 0.062 | 0.205 | 262 |
| | | | | *Kenya (CP1)* |
| *chr3:79,845 G>A (FIKK3)* | 7.2% | 1.000 | 1.000 | 1 |
| *chr3:96,476 C>T (PTP7)* | 23.6% | 0.118 | 0.936 | 1 |
| *chr3:124,242 T>G (CLAG3.2)* | 25.1% | 0.224 | 0.492 | 1 |
| *chr3:140,167 T>G (CLAG3.1)* | 22.8% | 0.229 | 0.444 | 1 |
| *chr3:79,845-140,167 (excluding the above)* | 2.3-12.3% | 0.040 | 0.330 | 558 |
| | | | | *Ghana* |
| *chr3:79,845 G>A (FIKK3)* | 5.9% | 1.000 | 1.000 | 1 |
| *chr3:96,476 C>T (PTP7)* | 14.9% | 0.515 | 0.872 | 1 |
| *chr3:140,167 T>G (CLAG3.1)* | 40.6% | 0.552 | 0.841 | 1 |
| *chr3:79,845-140,167 (excluding the above)* | 0.9-10.9% | 0.039 | 0.536 | 244 |

#### Supplementary Table 7

Linkage disequilibrium between a putative *Pfsa*+ locus in the gene PF3D7_0220500 (*Hyp2*) (chr2:825,357 T>C), the *Pfsa1*+ (chr2:631,190 T>A) and *Pfsa2*+ (chr2:814,288 C>T) loci and a mutation nearby to *Pfsa2*+ (chr2:814,297 C>T), shown separately for each dataset. For each pair, the minor allele frequency (MAF), the absolute Pearson's correlation coefficient (|*r*|) between genotypes, and the absolute D′ value is reported. The final row for each dataset shows the average statistic calculated using all sites in the specified range chr2:631,190-825,357) (excluding the four listed loci) whose MAF is within ±5% (and with a minimum MAF of 2.5%) of the MAF at the *Pfsa1*+ site in that dataset. "Number of sites" refers to the number of SNPs contributing to each computed value.

| *Position (range)* | *Minor allele frequency (range)* | *Absolute **Pearson's R** to chr2:825,357 T>C (Hyp2) (across range)* | *Absolute **D'** to chr2:825,357 T>C (Hyp2) (across range)* | *Number of sites* |
|---|---|---|---|---|
| | | | | *Gambia (GAMCC)* |
| *chr2:825,357 T>C (Hyp2)* | 7.7% | 1.000 | 1.000 | 1 |
| *chr2:631,190 T>A (Pfsa1)* | 9.1% | 0.452 | 0.483 | 1 |
| *chr2:814,288 C>T (Pfsa2)* | 0.0% | | | 1 |
| *chr2:814,297 C>T (0220300)* | 8.8% | 0.804 | 0.854 | 1 |
| *chr2:631,190-825,357 (excluding the above)* | 2.7-12.7% | 0.053 | 0.236 | 1853 |
| | | | | *Gambia (CP1)* |
| *chr2:825,357 T>C (Hyp2)* | 21.3% | 1.000 | 1.000 | 1 |
| *chr2:631,190 T>A (Pfsa1)* | 26.4% | 0.283 | 0.311 | 1 |
| *chr2:814,288 C>T (Pfsa2)* | 1.1% | 0.054 | 0.284 | 1 |
| *chr2:814,297 C>T (0220300)* | 18.3% | 0.621 | 0.722 | 1 |
| *chr2:631,190-825,357 (excluding the above)* | 16.3-26.3% | 0.065 | 0.129 | 553 |
| | | | | *Kenya (CP1)* |
| *chr2:825,357 T>C (Hyp2)* | 5.5% | 1.000 | 1.000 | 1 |
| *chr2:631,190 T>A (Pfsa1)* | 12.1% | 0.383 | 0.578 | 1 |
| *chr2:814,288 C>T (Pfsa2)* | 14.9% | 0.485 | 0.829 | 1 |
| *chr2:814,297 C>T (0220300)* | 0.0% | | | 1 |
| *chr2:631,190-825,357 (excluding the above)* | 2.5-10.5% | 0.034 | 0.193 | 1544 |
| | | | | *Ghana* |
| *chr2:825,357 T>C (Hyp2)* | 3.3% | 1.000 | 1.000 | 1 |
| *chr2:631,190 T>A (Pfsa1)* | 3.9% | 0.379 | 0.393 | 1 |
| *chr2:814,288 C>T (Pfsa2)* | 0.08% | 0.005 | 1.000 | 1 |
| *chr2:814,297 C>T (0220300)* | 1.3% | 0.560 | 0.878 | 1 |
| *chr2:631,190-825,357 (excluding the above)* | 2.5-8.3% | 0.035 | 0.242 | 152 |
